# Preregistration and Credibility of Clinical Trials^*^

**DOI:** 10.1101/2023.05.22.23290326

**Authors:** Christian Decker, Marco Ottaviani

## Abstract

Preregistration at public research registries is considered a promising solution to the credibility crisis in science, but empirical evidence of its actual benefit is limited. Guaranteeing research integrity is especially vital in clinical research, where human lives are at stake and investigators might suffer from financial pressure. This paper analyzes the distribution of p-values from pre-approval drug trials reported to *ClinicalTrials.gov*, the largest registry for research studies in human volunteers, conditional on the preregistration status. The z-score density of non-preregistered trials displays a significant upward discontinuity at the salient 5% threshold for statistical significance, indicative of p-hacking or selective reporting. The density of preregistered trials appears smooth at this threshold. With caliper tests, we establish that these differences between preregistered and non-preregistered trials are robust when conditioning on sponsor fixed effects and other design features commonly indicative of research integrity, such as blinding and data monitoring committees. Our results suggest that preregistration is a credible signal for the integrity of clinical trials, as far as it can be assessed with the currently available methods to detect p-hacking.

## I Introduction

Recently, there has been a growing interest in enhancing research transparency and credibility, both in social and life sciences.^1^ Randomized controlled trials (RCTs), as they are conducted commonly in clinical research and gain popularity in social sciences, are still considered the “gold standard” for empirical research. However, even such experiments have attracted skepticism regarding their credibility, as researchers have certain degrees of freedom about which hypotheses to test and report. This discretion may bias, intentionally or unintentionally, the set of published results, which may be the basis of important policy decisions. *Preregistration* of studies at public research registries such as *ClinicalTrials.gov* or the *American Economic Association*’s (AEA) RCT Registry can be a potential remedy for these concerns about credibility and integrity. Preregistration allows researchers to commit ex-ante to a study’s design, measured outcomes, and a protocol for statistical analysis and should shut down any tempting degrees of freedom during the research process.

Clinical studies with human volunteers, in particular, must comply with the highest ethical and scientific standards, as human lives may be directly at stake. At the same time, developing a new drug requires sizeable ex-ante investments,^2^ which can be typically recovered only if the trials are successful and authorities like the *U.S. Food and Drug Administration* (FDA) grant marketing approval. These financial considerations may lead to conflicts of interest and lure investigators into exploiting their degrees of freedom to beautify results or withhold unfavorable findings (Adda, Decker, and Ottaviani, 2020).

This paper analyzes the distribution of p-values from pre-approval drug trials reported to *ClinicalTrials.gov*, the largest registry for research studies in human volunteers, to assess if preregistration improves indicators of research credibility. In particular, we apply methods for detecting p-hacking and selective reporting to the samples of statistical results from preregistered and non-preregistered trials.^3^ P-hacking (i.e., intentionally or unconsciously exploring various ways of analyzing data and selectively reporting the ones that yield the best results) and selective reporting of results depending on their statistical significance are two of the main concerns of how researchers can exploit their degrees of freedom. According to the advocates of preregistration, non-preregistered research “will almost inevitably end in p-hacking,” and only committing ex-ante to an analysis plan through preregistration can prevent this temptation (Simmons, Nelson, and Simonsohn, 2021). However, there are also some critical voices arguing that the currently implemented preregistration systems are insufficient, still allow for researchers’ degrees of freedom, and give a false sense of improved credibility (Pham and Oh, 2021).^4^

Speaking to this debate, we find robust empirical evidence that the distribution of results from preregistered trials (defined as registered at *ClinicalTrials.gov* before the start of the trial) is significantly more credible than that from non-preregistered trials, as far as it can be assessed with the currently available methods for the detection of p-hacking.^5^

First, we apply density discontinuity tests (Cattaneo, Jansson, and Ma, 2020) to the densities of z-scores from preregistered and non-preregistered trials. For z-scores from primary outcomes of non-preregistered trials, we detect a substantial and statistically significant upward discontinuity at the prominent 5% threshold for statistical significance. Such a discontinuity is commonly interpreted as indicative of p-hacking to clear this salient significance hurdle or for selective non-reporting of results that did not achieve statistical significance. In contrast, the density of z-scores from primary outcomes of preregistered trials is smooth at this threshold.

The densities of z-scores related to lower-stake secondary outcomes, which are not consulted directly to evaluate a study’s success and are therefore less likely to be targeted by p-hacking, are smooth at the 5% threshold both for preregistered and non-preregistered trials.

Density discontinuity tests allow only for an unconditional analysis of the impact of preregistration on patterns indicative of p-hacking. However, we find that a trial’s preregistration status is substantially correlated with other design features which are commonly seen as signs of research integrity and credibility, such as blinding, the presence of an independent data monitoring committee, or the principal investigator (PI) not being on the sponsor’s payroll. Moreover, preregistration and complete and honest reporting of results may both be correlated with unobserved traits of the responsible researchers or unobservable incentives of sponsors. These issues raise the question of whether the differences detected by the density discontinuity tests are really driven by preregistration or instead by one of these other factors and show that a conditional analysis is required.

To address these concerns, we perform caliper tests that compare the share of significant results in a small discrete window of equal size below and above the significance threshold for different groups of trials (Gerber and Malhotra, 2008; Brodeur, Cook, and Heyes, 2020). Caliper tests can be embedded in a regression framework and therefore allow for the assessment of the effect of preregistration conditional on other covariates and fixed effects. Controlling for features like data monitoring committees, blinding, and independent PIs barely affects the magnitude and statistical significance of the estimated impact of preregistration. Moreover, none of these other features appears to have an effect on patterns indicative of p-hacking which is statistically significant or as large in magnitude as the impact of preregistration. Lastly, we estimate models with high-dimensional sponsor fixed effects to control for unobserved researcher traits or sponsor incentives. Even in these specifications, the differences between preregistered and non-preregistered trials are robust.

In summary, we do not find evidence of p-hacking in preregistered trials. In contrast, the density of z-scores from primary outcomes of non-preregistered trials exhibits patterns indicative of p-hacking or selective reporting. The difference between preregistered and non-preregistered trials regarding these indicators appears robust to conditioning on trials’ other design characteristics and sponsor fixed effects capturing unobserved researcher traits and sponsor incentives. We conclude that preregistration under the current regime of regulations is indeed a reliable signal for research integrity and credibility as far as it can be evaluated with the currently available tools for detecting p-hacking and selective reporting. While we cannot provide bullet-proof evidence from exogenous or quasi-exogenous variation in the trials’ preregistration status, our results with high-dimensional sponsor fixed effects may be indicative of preregistration having a causal impact on the integrity and credibility of reported results.

This paper complements a series of other studies about the effects of preregistration in different contexts. So far, only a limited impact of preregistration on measures of research integrity could be documented empirically. Brodeur et al. (2022) find that preregistration does not affect patterns indicative of p-hacking or selective reporting for randomized controlled trials published in leading economics journals. However, studies with a complete pre-analysis plan appear significantly less p-hacked. Abrams, Libgober, and List (2023) present evidence that the uptake and quality of registrations in the AEA Registry and *ClinicalTrials.gov* are relatively poor and find that preregistration at the AEA registry does not impact indications of p-hacking in a randomly selected sample of economics RCTs. Moreover, they explore policies that could increase registry effectiveness in a theoretical model.^6^ Fang, Gordon, and Humphreys (2015) find no evidence that preregistration requirements for publication in medical journals had any impact on publication bias. Oostrom (2022) shows that preregistration on *ClinicalTrials.gov* mitigates sponsorship bias for trials on antidepressants and antipsychotics.

The remainder of this paper is organized as follows. Section II introduces the context of the *ClinicalTrials.gov* registry, discusses the composition of our sample of p-values, and elaborates on preregistration and reporting requirements. Section III presents results from density discontinuity tests for z-scores from preregistered and non-preregistered trials. Section III presents results from caliper tests, which allow to condition on other trial characteristics and fixed effects. Finally, Section V concludes. Supplementary results and robustness checks are gathered in an Online Appendix.

## II Background and Data

### The *ClinicalTrials.gov* Registry

*ClinicalTrials.gov*, maintained by the *National Library of Medicine* (NLM) at the *National Institutes of Health* (NIH) in collaboration with the FDA, is the largest online registry of clinical research studies in human volunteers. The registry was established in 2000 to increase transparency in clinical research (Zarin and Tse, 2008). In 2010, the *Clinical Trials Transformation Initiative*, a partnership of the FDA and Duke University, launched the *Database for Aggregate Analysis of ClinicalTrials.gov* (AACT), allowing for free bulk download of all the data contained in the *ClinicalTrials.gov* registry (Tasneem et al., 2012). The daily updated database contains information on all aspects of registered trials, including timing, interventions, sponsors, outcome measures, and results.

The results in the database pertain to a wide range of different diseases, interventions, study designs, and statistical procedures. As we aim to analyze the largest possible portion of the overall data, we concentrate on p-values, the only measure reported uniformly and comparably for many trials, independent of their characteristics and the statistical method used for the analysis.

### Sample

This paper is based on the AACT snapshot from February 18, 2023. Figure 1 visualizes the construction of our sample. We focus on completed or terminated pre-approval (phase II, phase III, combined phase II/III) interventional (as opposed to observational) studies on drugs (as opposed to medical devices and others) that report at least one exact p-value for a statistical test on a primary outcome of the trial aiming to establish the superiority of the treatment (as opposed to non-inferiority or equivalence).^7^ The resulting sample contains 10,120 p-values from tests performed on primary outcomes of 4,810 trials and 54,337 p-values from tests performed on secondary outcomes.^8^ Further summary statistics are discussed in Online Appendix A.

**Figure 1:**
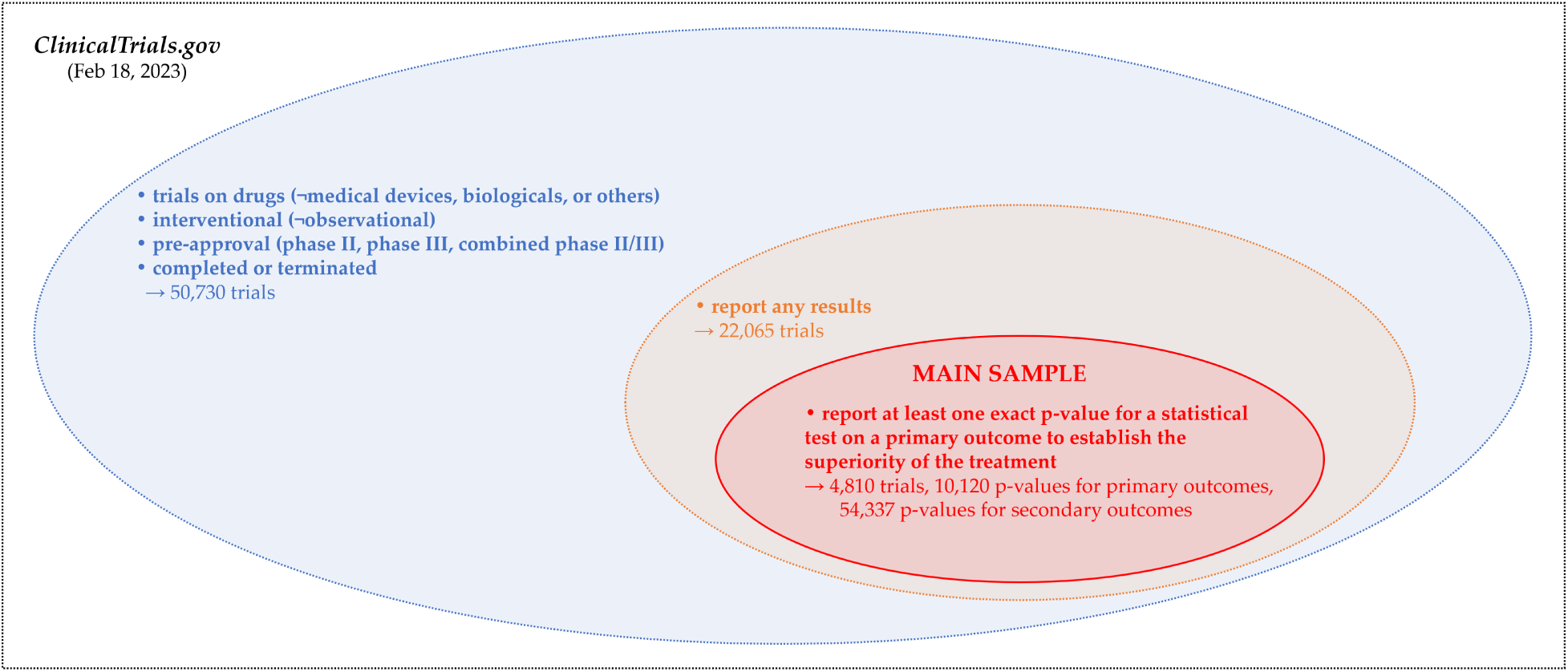
Sample Composition

### p-z Transformation

We transform reported p-values to z-scores by supposing that these p-values originate from a two-sided Z-test of the null hypothesis that the drug has the same effect as the comparison.^9^ Under the null hypothesis, these z-scores are Gaussian, so we have the one-to-one correspondence

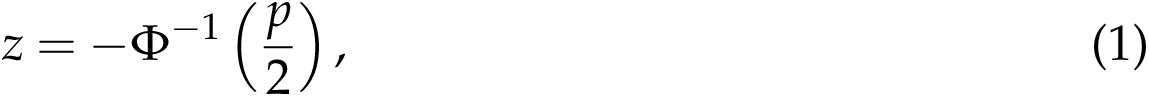

where *z* is the absolute value of the test statistic, and Φ*^−^*^1^ is the inverse of the standard normal cumulative distribution function. While the density of p-values is highly convex around the 5% significance threshold, the density of corresponding z-scores is closer to linear in this range, making it easier to identify a discontinuity at the threshold. Moreover, the density of z-scores is more convenient for graphical analysis; to make potential irregularities at the significance threshold visible, we do not need to zoom in to a small window, but we can plot the entire distribution.

### Registration and Reporting Requirements

Section 801 of the *FDA Amendments Act* (FDAAA), initially passed in 2007 and in effect since April 2017, describes a set of criteria under which the responsible party (i.e., the trial’s sponsor or principal investigator) is prescribed to^10^

- register a trial at *ClinicalTrials.gov* no later than 21 days after the start date,^11^ and
- submit results of a trial at *ClinicalTrials.gov* no later than twelve months after the primary completion date.^12^

### Preregistration

The information that the initial registration needs to contain, according to the FDAAA, includes descriptive information about the trial (e.g., title, design, primary outcome measures, timeline), recruitment information (e.g., eligibility criteria), and location and contact information (e.g., name of the sponsor, facility information). A complete pre-analysis plan, as nowadays seen often for RCTs in economics, is neither required nor commonly found on *ClinicalTrials.gov*. The registration can be updated and modified at any time, and investigators do so frequently by gradually adding more details about the study protocol and progress. The main page of the registry shows the information of the latest submission. The entire history of modifications is stored and accessible with a few clicks, but the information of earlier submissions (and, in particular, the initial submission) is not displayed very prominently. This design reflects that the primary intention of the registry has not been to provide a platform for preregistration of trials but a platform that collects information from *all* trials, including those that were “not successful” and would not lead to publications in academic journals, and makes them easily accessible.

Strictly speaking, there is no legal preregistration requirement *before* the start of a trial, but the first registration has to occur within three weeks after the trial has started. Independent of the FDA regulation, already since 2005, the *International Committee of Medical Journal Editors* (ICMJE) has required preregistration “at or before the onset of patient enrollment” at a public trials registry such as *ClinicalTrials.gov* (or one of the smaller registries of other countries) as a condition to be considered for publication in many of the major medical journals (De Angelis et al., 2004).

For our analysis, we follow the ICMJE and define a trial as preregistered if the first registration at *ClinicalTrials.gov* was submitted at or before the trial’s start date. We define a trial as non-preregistered if the first submission to the registry was after the start date. Figure 2 visualizes the timeline of the registration and reporting process. Note that the initial submission of a study may occur even after the completion of the trial.

**Figure 2:**
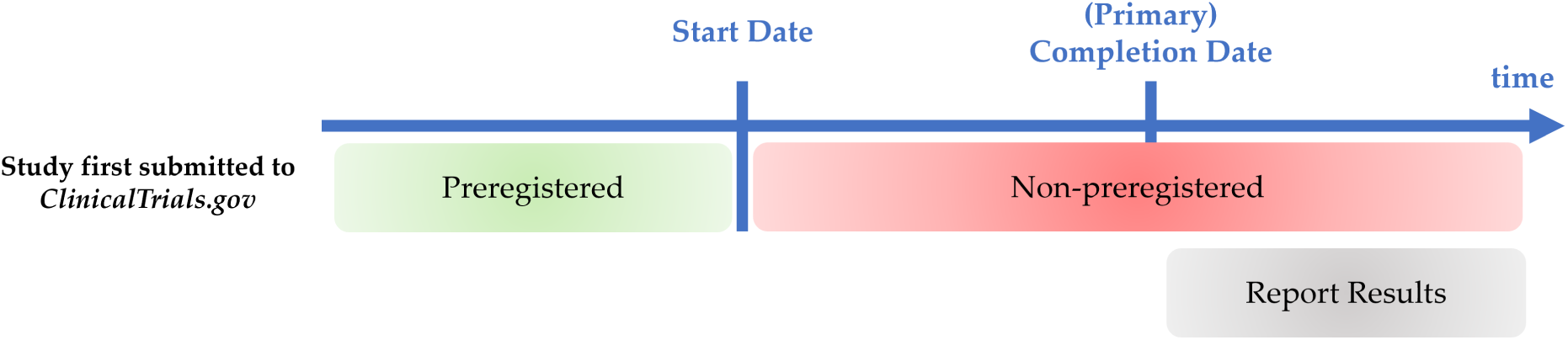
Timeline of Registration and Reporting

Figure 3 shows how the number of trials in the registry and the share of preregistered trials have evolved over time by the start year of the trials. The bars in the background display the number of trials in the registry, which fall in the different groups defined in Figure 1 and share the same color coding. The registry gained importance with the first introduction of the FDAAA in 2007. Since then, 3,000 to 4,000 trials per year were submitted until 2019. In 2020 and 2021, this number increased to almost 5,000. The numbers for 2022 and 2023 are lower as many trials that started or are about to start in these years may not have been registered yet. The share of preregistered trials (according to our definition discussed in the previous paragraph), depicted by the green line, has continuously increased up to a plateau of about 80%. The strong uptick to almost 1 in the last two years is mainly mechanical, as many of the non-preregistered trials started in these years may not have been reported to the registry yet.

**Figure 3:**
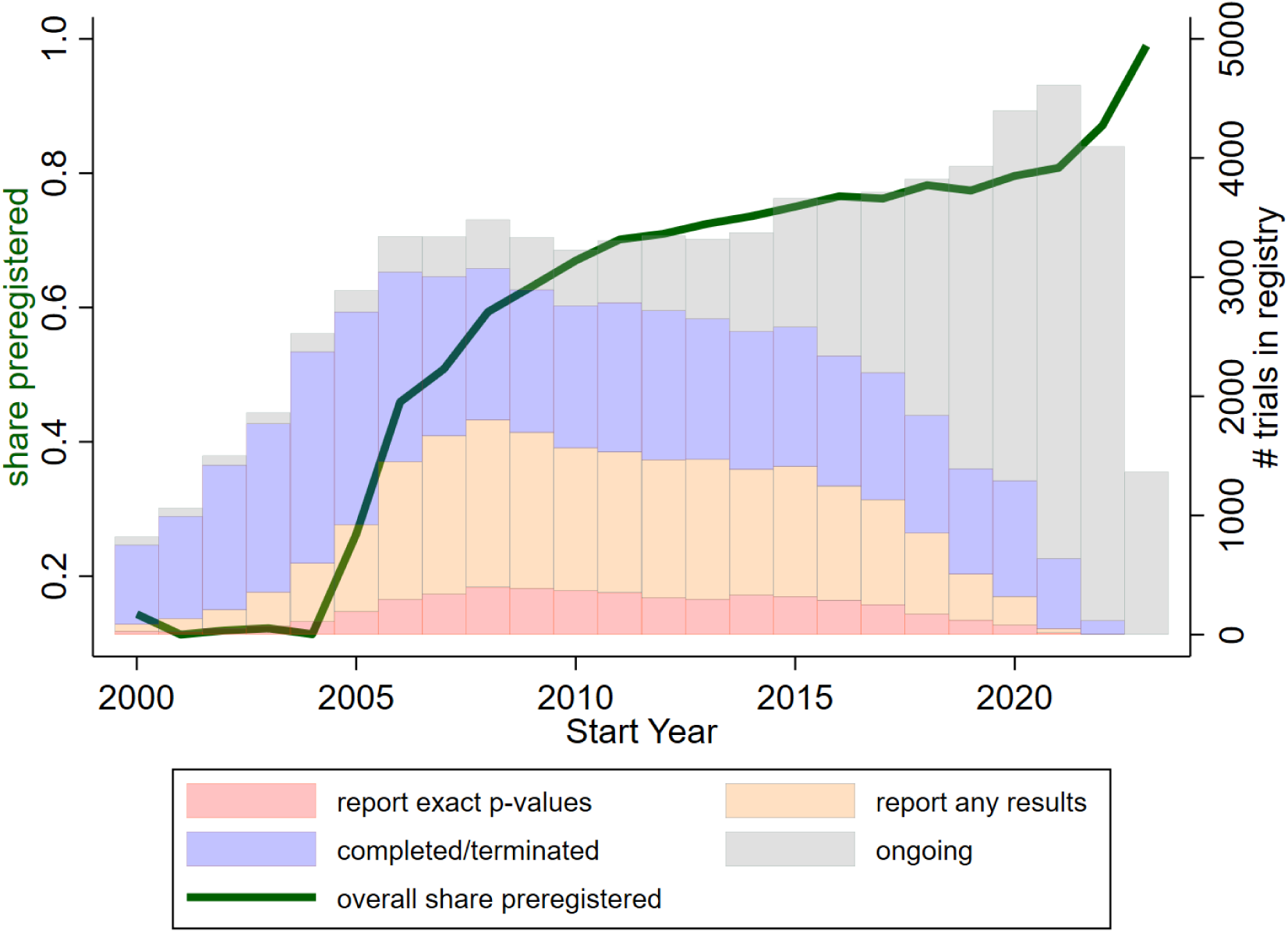
Time Trends *Notes:* The bars in the background (right y-axis) show the number of trials in the registry by start year. The color coding corresponds to the groups defined in Figure 1. The gray bars refer to ongoing trials that (except completion) meet all the other criteria to be included in our sample. The green line (left y-axis) shows the share of preregistered trials (defined as first submitted to the registry at or before the start date) among all trials (blue ellipse in Figure 1 plus the ongoing trials) by start year.

The preregistration status of trials is correlated with other important characteristics and design features. Table 1 assesses the balancedness of such other trial characteristics between preregistered and non-preregistered trials in our main sample.^13^ The first set of characteristics we consider are design features that, like preregistration, are commonly regarded as good practice and a sign of integrity and credibility. These include the presence of an independent data-monitoring committee, blinding of subjects, caregivers, investigators, and outcome assessors, and the principal investigator not being an employee of the study’s sponsor. All of these binary variables are coded with value one if the design is ”superior” in terms of integrity.^14^ Except for the masking of the caregiver, preregistered trials are significantly more likely to feature these other superior design elements, with differences ranging between 3 and 11 percentage points. Moreover, preregistered trials are significantly more likely to involve a placebo control and to be industry-sponsored, as well as less likely to be in phase III (instead of phase II or phase II/III combined). There do not seem to be substantial differences in the number of enrolled subjects and the number of reported outcomes.

**Table 1:** Comparing Characteristics of Non-Preregistered and Preregistered Trials in Main Sample

|  | (1)<br>non-preregistered | (2)<br>preregistered | (3)<br>difference (2)–(1) |
| --- | --- | --- | --- |
| data monitoring committee | 0.464<br>(0.499) | 0.575<br>(0.494) | 0.111***<br>(0.018) |
| subject masked | 0.711<br>(0.453) | 0.768<br>(0.422) | 0.057***<br>(0.014) |
| caregiver masked | 0.420<br>(0.494) | 0.444<br>(0.497) | 0.024<br>(0.017) |
| investigator masked | 0.697<br>(0.460) | 0.751<br>(0.433) | 0.053***<br>(0.015) |
| outcomes assessor masked | 0.398<br>(0.490) | 0.431<br>(0.495) | 0.033**<br>(0.016) |
| mask folds | 2.226<br>(1.576) | 2.394<br>(1.505) | 0.168***<br>(0.051) |
| PI no employee of sponsor | 0.878<br>(0.327) | 0.914<br>(0.281) | 0.036***<br>(0.010) |
| enrollment | 632.6<br>(1,764.7) | 549<br>(1,573.6) | -83.6<br>(54.0) |
| placebo-controlled | 0.634<br>(0.482) | 0.714<br>(0.452) | 0.079***<br>(0.015) |
| industry-sponsored | 0.684<br>(0.465) | 0.763<br>(0.425) | 0.079***<br>(0.014) |
| phase III | 0.551<br>(0.498) | 0.508<br>(0.500) | -0.044***<br>(0.017) |
| # exact p-values from primary outcomes | 2.108<br>(2.194) | 2.103<br>(2.288) | -0.005<br>(0.075) |
| # exact p-values from secondary outcomes | 9.483<br>(37.684) | 11.904<br>(54.005) | 2.421<br>(1.677) |
| Observations | 1,206 | 3,604 | 4,810 |

*Notes:* The first two columns show the means (and, in parentheses, standard deviations) of the variables indicated in the respective row for non-preregistered and preregistered trials in our main sample (red ellipse in Figure 1). Column 3 shows the differences between the first two columns with standard errors in parentheses; significance levels indicated by the stars: \* $p < 0.1$ , \*\* $p < 0.05$ , \*\*\* $p < 0.01$ . The variable “mask folds” adds up the four dummy variables in the four rows above.

Table 1 shows the importance of controlling for other design features when assessing the impact of preregistration on the distribution of reported p-values, as we set out to do with the caliper tests in Section IV.

### Reporting of Results

Trial results that are supposed to be reported to *ClinicalTrials.gov* according to the FDAAA include the participant flow, demographic and baseline characteristics of different treatment groups, adverse event information, and outcomes and statistical analyses for each primary and secondary outcome measure. The latter may contain the p-values we focus on for our analysis. These result data are usually submitted after the primary completion of the trial (see Figure 2).

Compliance with the FDAAA regulations is still poor, especially regarding reporting results to the registry.^15^ Of the group of trials we look at, only 43.5% reported any results to the registry at all, and 9.5% reported at least one exact p-value to be included in our sample (see Figure 1).^16^ The time trend of reporting rates can be read in Figure 3.

A natural limitation of our study is that we can only analyze the distribution of p-values for those trials that actually do report results to the registry. To assess the representativeness of our sample, Table 2 compares the characteristics and design of trials that report at least one exact p-value (column 1, our main sample, red ellipse in Figure 1), trials that report some results but no exact p-values (column 2, orange but not red ellipse in Figure 1), and trials that do not report any results (column 3, blue but not orange ellipse in Figure 1).

**Table 2:** Comparing Characteristics of Trials that Report P-Values, Trials that Report Results, and Trials that Do not Report Results

|  | (1)<br>exact p-values<br>provided | (2)<br>results provided,<br>but no exact p-values | (3)<br>no results<br>provided | (4)<br>difference<br>(1)–(2) | (5)<br>difference<br>(1)–(3) | (6)<br>difference<br>(2)–(3) |
| --- | --- | --- | --- | --- | --- | --- |
| preregistered | 0.749<br>(0.433) | 0.669<br>(0.471) | 0.435<br>(0.496) | 0.080***<br>(0.008) | 0.314***<br>(0.008) | 0.234***<br>(0.005) |
| data monitoring committee | 0.549<br>(0.498) | 0.495<br>(0.500) | 0.469<br>(0.499) | 0.053***<br>(0.009) | 0.079***<br>(0.008) | 0.026***<br>(0.005) |
| subject masked | 0.754<br>(0.431) | 0.421<br>(0.494) | 0.388<br>(0.487) | 0.333***<br>(0.008) | 0.366***<br>(0.007) | 0.034***<br>(0.005) |
| caregiver masked | 0.438<br>(0.496) | 0.238<br>(0.426) | 0.229<br>(0.420) | 0.199***<br>(0.007) | 0.208***<br>(0.007) | 0.009**<br>(0.004) |
| investigator masked | 0.737<br>(0.440) | 0.405<br>(0.491) | 0.363<br>(0.481) | 0.332***<br>(0.008) | 0.374***<br>(0.007) | 0.043***<br>(0.005) |
| outcome assessor masked | 0.423<br>(0.494) | 0.237<br>(0.425) | 0.224<br>(0.417) | 0.187***<br>(0.007) | 0.199***<br>(0.007) | 0.013***<br>(0.004) |
| mask folds | 2.352<br>(1.524) | 1.301<br>(1.604) | 1.203<br>(1.583) | 1.051***<br>(0.026) | 1.149***<br>(0.025) | 0.098***<br>(0.015) |
| PI no employee of sponsor | 0.905<br>(0.294) | 0.781<br>(0.414) |  | 0.124***<br>(0.006) |  |  |
| enrollment | 569.9<br>(1,623.9) | 242.1<br>(999.2) | 245.0<br>(978.0) | 327.9***<br>(19.0) | 324.9***<br>(17.2) | -2.9<br>(9.6) |
| placebo-controlled | 0.694<br>(0.461) | 0.350<br>(0.477) | 0.289<br>(0.453) | 0.344***<br>(0.008) | 0.404***<br>(0.007) | 0.060***<br>(0.004) |
| industry-sponsored | 0.743<br>(0.437) | 0.572<br>(0.495) | 0.466<br>(0.499) | 0.172***<br>(0.008) | 0.278***<br>(0.008) | 0.106***<br>(0.005) |
| phase III | 0.519<br>(0.500) | 0.368<br>(0.482) | 0.389<br>(0.488) | 0.151***<br>(0.008) | 0.130***<br>(0.008) | -0.021***<br>(0.005) |
| Observations | 4,810 | 17,255 | 28,665 | 22,065 | 33,475 | 45,920 |

*Notes:* The first three columns show the means (and, in parentheses, standard deviations) of the variables indicated in the respective row for the trials in the different samples. Column 1 refers to the main sample of trials that report exact p-values (red ellipse in Figure 1). Column 2 refers to trials that provide results but no exact p-values (orange ellipse but not red ellipse in Figure 1). Column 3 refers to applicable trials that do not report results (blue ellipse but not orange ellipse in Figure 1). Columns 4-6 show pairwise differences of the first three columns with standard errors in parentheses; significance levels indicated by the stars: \* $p < 0.1$ , \*\* $p < 0.05$ , \*\*\* $p < 0.01$ . The variable “mask folds” adds up the four dummy variables in the four rows above. Information about the PI’s employment is unavailable for trials that do not report results.

The trials in our main sample are 8 percentage points more likely to be preregistered than trials that report results but no exact p-value and 31 percentage points more likely to be preregistered than trials that do not report any results. Regarding the other superior design characteristics (data monitoring committee, blinding, independent PI), trials in our sample appear substantially better than trials that report results but no exact p-values. However, the differences between the trials that report results but no exact p-values and those trials that do not report any results appear less pronounced. Moreover, trials in our sample enroll, on average, more than twice the number of participants and are substantially more likely to involve a placebo control, be industry-sponsored, and be in phase III.

In summary, it seems that our sample of p-values stems from the group of larger-scale trials that do best regarding research integrity and transparency standards. As such, we would expect any concerns of p-hacking we detect for this sample to be even more pronounced for the other groups of trials, for which we do not have the chance to perform such an analysis.

## III Density Discontinuity Tests

### Method

In the absence of selective reporting or manipulation of results, the density of z-scores *f* (*z*) is continuous under a wide range of assumptions (Andrews and Kasy, 2019). Therefore, an intuitive and powerful way to test for manipulation to clear a certain significance threshold *c* is to test for a discontinuity in the density of z-scores at this threshold (Adda, Decker, and Ottaviani, 2020; Elliott, Kudrin, and Wüthrich, 2022*a*), formally

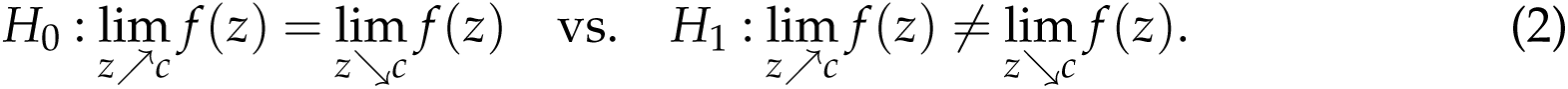

We implement the testing procedure proposed by Cattaneo, Jansson, and Ma (2020), an advancement of the McCrary (2008) test. This procedure builds on local polynomial density estimators with bias correction at the boundary *c*. The main advantages of this approach over other methods to detect p-hacking are that it avoids pre-binning of the sample and features an entirely data-driven bandwidth selection, allowing for possibly different bandwidths to the left and right of the testing threshold. Moreover, it does not restrict the sample of z-scores to a window around the testing threshold ex-ante but potentially exploits information of the entire distribution.

As an interpretable measure for the size of a discontinuity, we introduce

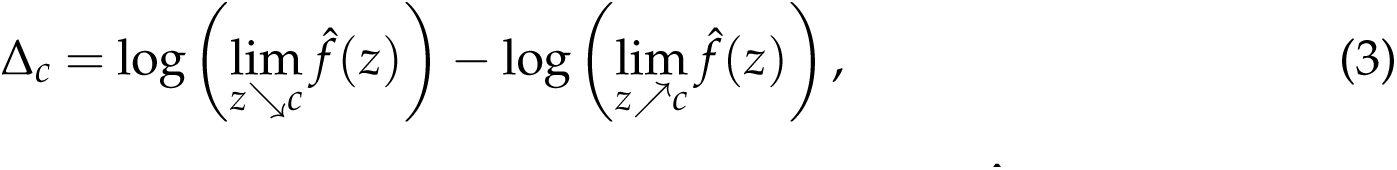

the log-difference in the limits of the bias-corrected density estimate and the left of *c*.

Our main analysis focuses on the most prominent threshold for statistical significance at the 95% confidence level, corresponding to a p-value smaller than 0.05 or a z-score large than 1.96.

### Tests for Primary Outcomes

Figure 4 shows histograms and density estimates with a discontinuity test at *z* = 1.96 (the threshold for statistical significance at the 5% level) for the distribution of z-scores from tests on *primary* trial outcomes. Primary outcomes are the endpoints that are most relevant to answer the specific research question of a trial; usually, these are patient-centered measures such as quality of life and survival. The trials in our main sample report, on average, 2.1 exact p-values for tests related to primary outcomes. Since a statistically significant impact of the tested interventions on these primary outcomes is often pivotal for the evaluation of a trial’s success (for instance, concerning the marketing approval decision of regulatory authorities such as the FDA or publication in medical journals), we would expect that any potential manipulation of results or selective reporting would focus on these primary outcomes.

**Figure 4:**
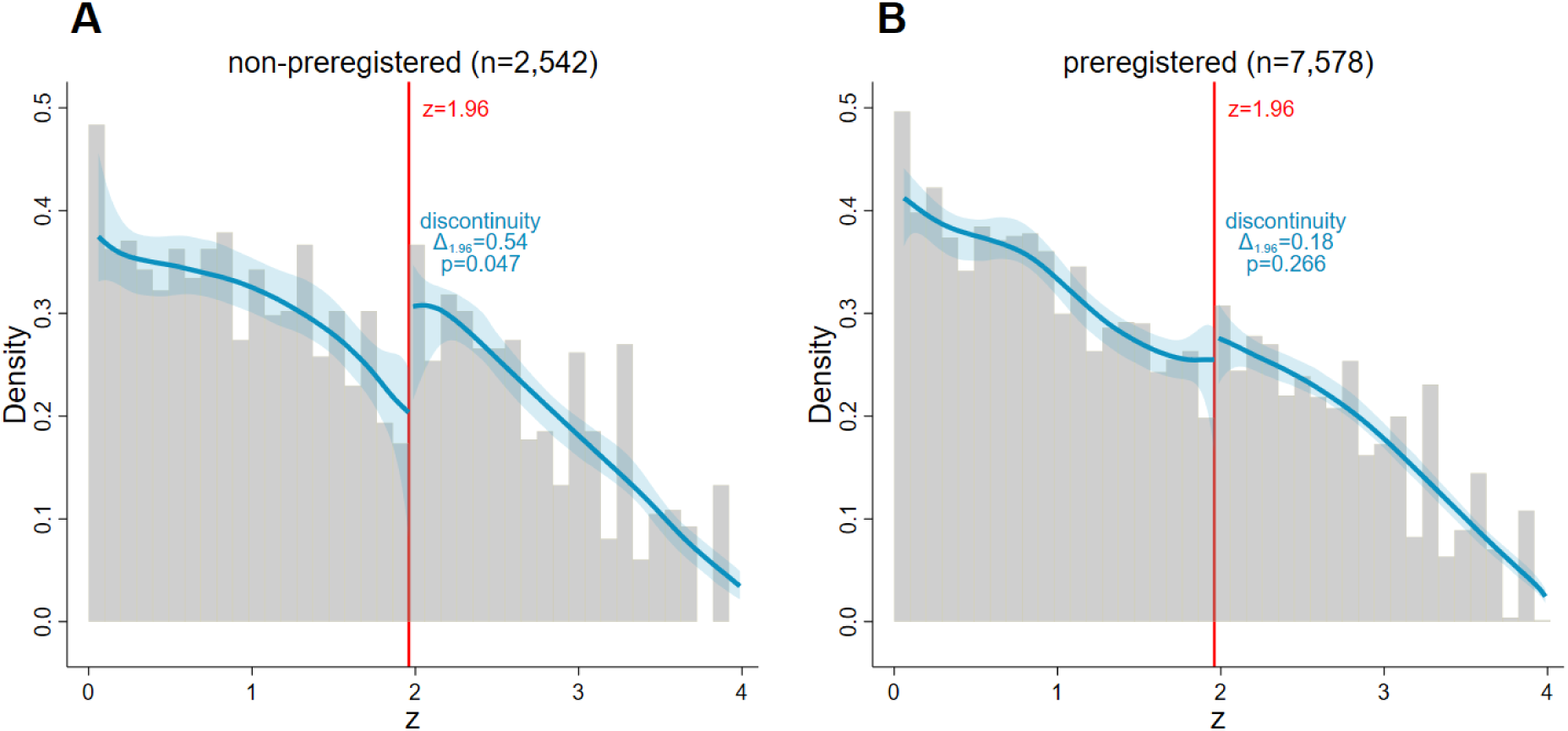
Density Discontinuity Tests at *z* = 1.96 (Primary Outcomes) *Notes:* The graphs show histograms (grey) and density estimates (blue) for the distribution of z-scores from tests on primary outcomes of non-preregistered (panel A) and preregistered trials (panel B). The shaded blue areas are 95% confidence bands for the density estimates, and the vertical red lines at 1.96 correspond to the threshold for statistical significance at the 0.05 level. The local polynomial density estimators proposed by Cattaneo, Jansson, and Ma (2020) are used. We allow for a discontinuity at 1.96 and present the log-difference measure Δ_1.96_ as defined in (equation 3 and the p-values of the discontinuity test (equation 2). Note that potential discrepancies between Δ_1.96_ and the differences that can be read off the plotted densities are due to the bias correction in the testing procedure.

The density discontinuity test for results from non-preregistered trials in panel A finds a statistically significant discontinuity at *z* = 1.96, indicative of some degree of p-hacking to clear the significance threshold or selective reporting of results based on statistical significance. The log-difference measure Δ_1.96_ suggests that the bias-corrected estimated density is 54% larger on the right of the cutoff than on the left.

Panel B, instead, considers preregistered trials. The estimated density is almost smooth at the significance threshold, and our test does not detect a statistically significant discontinuity. We conclude that preregistration can be seen as a reliable signal for the credibility of clinical trials, at least insofar as we do not find any evidence for p-hacking for reported results from trials preregistered at *ClinicalTrials.gov*.

### Tests for Secondary Outcomes

Besides the primary outcomes, which are the main endpoints of the studies, most trials also document statistical results for an array of *secondary outcomes*. Secondary outcomes are outcomes of lesser importance than primary outcomes, which are monitored additionally to help interpret the results of primary outcomes. As such, we would not expect secondary outcomes to be the target of p-hacking. Of the trials in our main sample, 67% report at least one exact p-value for a statistical test of a secondary outcome; conditional on reporting exact p-values for secondary outcomes, the average number of reported results is 16.9.

Figure 5 shows histograms and density estimates for the distribution of z-scores from secondary outcomes. Both for non-preregistered trials (panel A) and preregistered trials (panel B), the estimated densities are very smooth around the significance threshold at *z* = 1.96, and we do not find any evidence for p-hacking.

**Figure 5:**
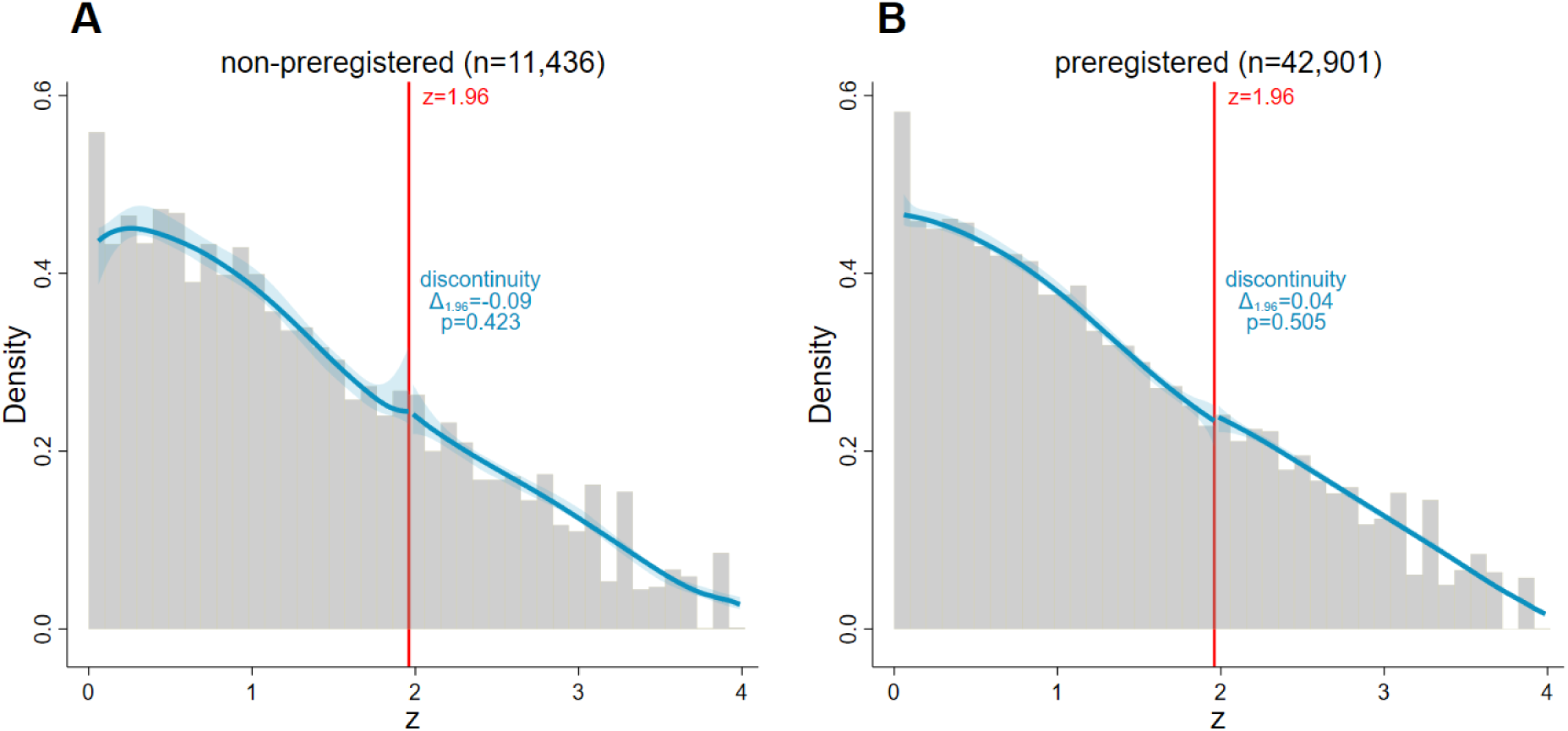
Density Discontinuity Tests at *z* = 1.96 (Secondary Outcomes) *Notes:* The graphs show histograms (grey) and density estimates (blue) for the distribution of z-scores from tests on secondary outcomes of non-preregistered (panel A) and preregistered trials (panel B). The shaded blue areas are 95% confidence bands for the density estimates, and the vertical red lines at 1.96 correspond to the threshold for statistical significance at the 0.05 level. The local polynomial density estimators proposed by Cattaneo, Jansson, and Ma (2020) are used. We allow for a discontinuity at 1.96 and present the log-difference measure Δ_1.96_ as defined in equation 3 and the p-values of the discontinuity test (equation 2). Note that potential discrepancies between Δ_1.96_ and the differences that can be read off the plotted densities are due to the bias correction in the testing procedure.

### Additional Results and Robustness Checks

In Online Appendix B, we show that the discontinuity for non-preregistered trials in Figure 4 is mainly driven by trials with a substantial reporting delay (*>*10% of trial duration) with more opportunity for p-hacking than only slightly delayed trials. Moreover, we show that density discontinuity tests do not detect irregularities at other significance thresholds (0.1, 0.01, 0.001) and that other design characteristics of trials are less correlated to a discontinuity at the significance threshold than the preregistration status. Also, we establish the robustness of our main result with respect to transforming p-values to one-sided instead of two-sided test statistics and de-rounding p-values. Finally, we apply the battery of alternative tests to detect p-hacking proposed by Elliott, Kudrin, and Wüthrich (2022*a*) to our data.

## IV Caliper Tests

### The Need of a Conditional Analysis

Recall from the discussion around Table 1 in Section II that the preregistration status of a trial is substantially correlated with other design features of the trial, which are commonly considered indicative of the integrity and credibility of research and might restrict the opportunity to p-hack. This begs the question if preregistration is really the decisive factor for the patterns we documented with the density discontinuity tests. To isolate the impact of preregistration on the distribution of reported p-values, these other characteristics need to be controlled for.

More generally, to obtain the causal effect of preregistration on p-hacking and selective reporting, we would need to compare the reported outcomes of a preregistered trial to those of an exactly identical hypothetical counterfactual trial by the same researchers that is non-preregistered, and vice-versa. Running an experiment to randomly assign trials to preregister or not preregister is not feasible in this context. Moreover, legal registration requirements are enforced too poorly to provide a natural experiment with plausible quasi-random variation in the preregistration status. Therefore, the best we can do to get close to a causal estimate is to control for observable trial characteristics and sponsor fixed effects as well as possible.

Density discontinuity tests require a discrete sample split to assess the impact of a particular variable on the discontinuity. They do not allow conditioning of the analysis on other variables, if not by splitting the sample even further. As the method is also quite demanding regarding the sample size required for robust results, slicing the sample in this way would not be practical. Therefore, we rely on caliper tests to assess the impact of preregistration conditional on other trial characteristics and fixed effects.

### Method

The basic idea of the caliper test, first introduced by Gerber and Malhotra (2008), is to compare the number of z-scores in a narrow, equally-sized window above and below the significance threshold. If significantly more results fall in the window above the threshold, this is considered evidence for p-hacking or selective reporting based on the significance level. As noted by Brodeur, Cook, and Heyes (2020), caliper tests allow to asses simultaneously the impact of several variables and fixed effects on the share of significant results within the specified window in a multivariate regression framework.

For a significance threshold *c* and a bandwidth *h*, we estimate for the sample of all z-scores in the window [*c − h*, *c* + *h*] the regression

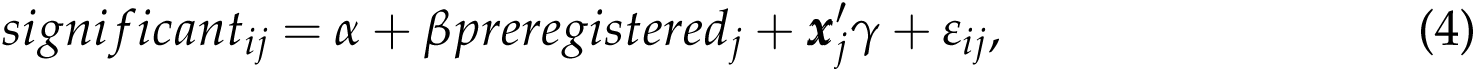

where *significant_ij_* is a dummy indicating if z-score *i* of trial *j* is above the significance threshold *c*, *preregistered_j_*is a dummy indicating if trial *j* was preregistered, ***x****_j_*is a vector of other characteristics of trial *j* (possibly including fixed effects), and *ε_ij_*is an error term. We use a linear probability model such that we can estimate specifications with high-dimensional sponsor fixed effects. In Online Appendix C, we show that the results without high-dimensional fixed effects are robust in probit and logit models. As the density discontinuity tests detected a discontinuity only at the 5% significance threshold, we focus our analysis on the cutoff *c* = 1.96.

### Choosing the Bandwidth

As discussed above, without manipulation, the density of z-scores is continuous. Therefore, a discontinuity at the significance threshold can be assigned to some sort of manipulation with certainty. With the caliper tests, we move away from investigating a discontinuity in the limit to a discrete window of scores below and above the threshold.^17^ Differences in the number of scores in two discrete bins may not necessarily be due to manipulation but may arise naturally as the density of z-scores is not flat but has a nonzero slope at the threshold. Elliott, Kudrin, and Wüthrich (2021) note that for certain distributions of “true” underlying effects, even upward-sloping portions of the z-density cannot be ruled out.

Moreover, in the proposed regression framework, we look at differences in caliper tests between groups of trials. These different groups of trials may not have the same underlying distributions of “true” effects, and their z-score densities may not have the same slope around the threshold. These natural differences may influence the estimated regression coefficients, which may pick up not only differences in a discontinuity at the threshold but, in part, also differences in the distributions of underlying “true” effects.

Intuitively, both the concerns that the caliper tests pick up the slope of the density instead of a discontinuity and that the regression coefficients are “contaminated” by differences in the underlying distributions of “true” effects become larger as the bandwidth *h* increases. However, smaller bandwidths will lead to smaller sample sizes and less statistical power. Therefore, a wise bandwidth choice is vital for the reliability of the caliper tests.

Led by the results of the density discontinuity tests and visual inspection of the histograms of the raw data in Figure 4, we set *h* = 0.2 as bandwidth for our main analysis. This corresponds roughly to two bins below and two bins above the significance threshold in the histograms. Visually, for non-preregistered trials, mass appears to be missing specifically in the two bins below the threshold, whereas the two bins above the threshold evidently feature an excess number of scores. Moreover, it might be challenging to inflate the z-score subtly by a margin of more than 0.2. Further down, we will show the estimates of the main coefficient of interest for alternative bandwidths.

In summary, the caliper tests have the crucial advantage that they can be embedded in a regression framework and can provide estimates for the impact of preregistration conditional on other trial characteristics. On the flip side, the results need to be interpreted carefully because the regression may pick up not only manipulation but also other (innocent) factors. We calibrate the caliper tests such that the baseline results align with the more precise density discontinuity tests, and we can be confident that the differences the caliper tests detect indeed indicate a different prevalence of p-hacking or selective reporting and are not driven by other factors.

### Baseline Results

Table 3 shows the results of the caliper test regression 4. Column 1 considers the baseline case without any further controls. In the group of non-preregistered trials, 63.3% of the scores that fall in the window [1.96 *±* 0.2] are significant. Confirming the results of the density discontinuity tests, this share is 8.6 percentage points lower for preregistered trials, a statistically significant difference at the 5% level.

**Table 3:**
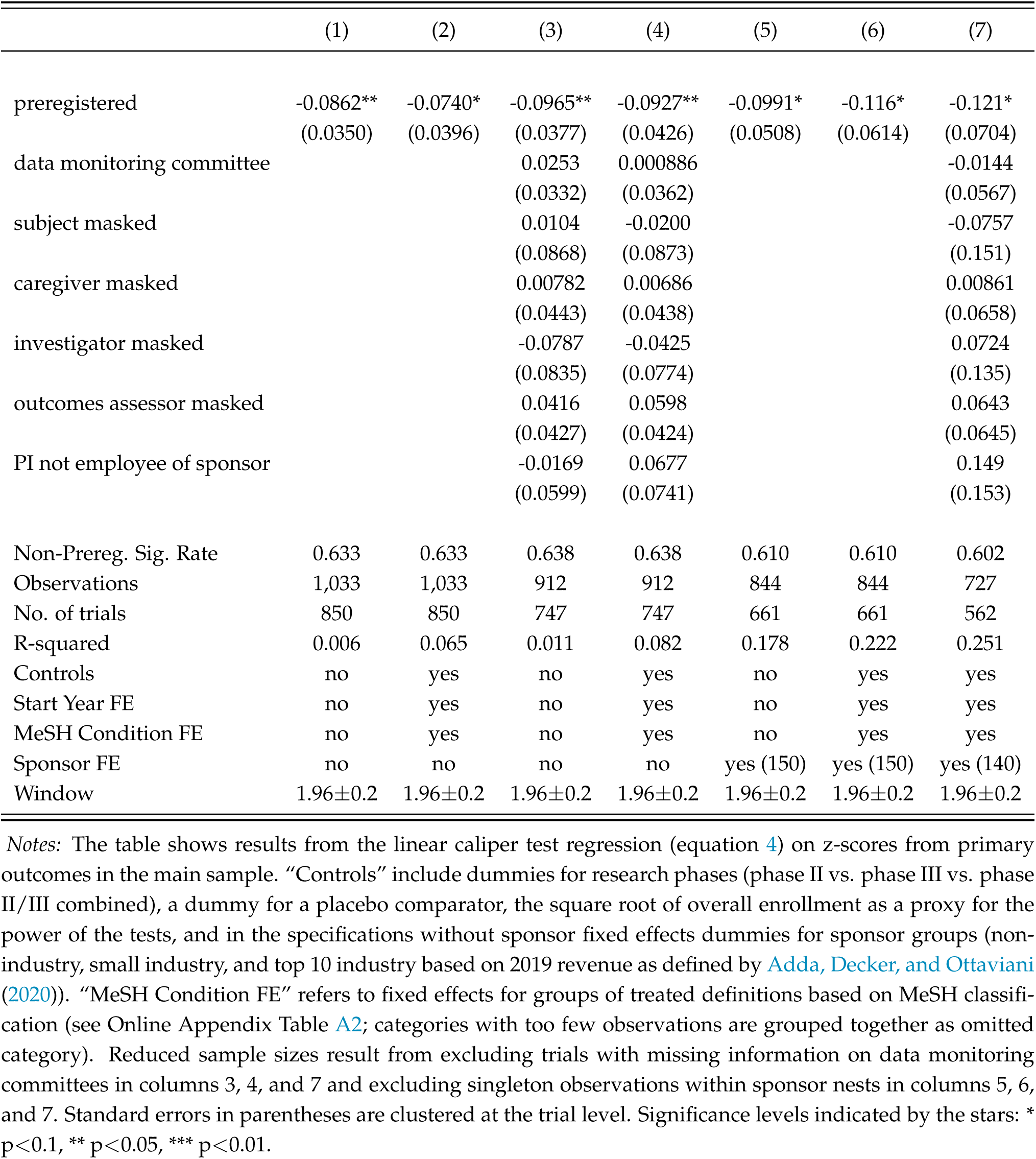
Caliper Tests for *z ∈*1.96*±*0.2 (Primary Outcomes, LPM)

In column 2, we start to control for other trial characteristics; these are dummies for sponsor groups (large industry, small industry, non-industry), phases (phase II, phase III, phase II/III combined), and the trial involving a placebo control, as well as the square root of the number of enrolled participants as a proxy for the power of the statistical tests. Moreover, we include fixed effects for the start years of the trial to control flexibly for any time-trend, and fixed effects for the treated conditions based on *Medical Subject Headings* (MeSH) classification. The resulting estimate of the difference between preregistered and non-preregistered trials and its precision differ only marginally from the baseline in column 1.

### Other Design Features

Columns 3 and 4 repeat the specifications of the first two columns but control additionally for the other superior design characteristics that are, as shown in Table 1, positively correlated with a trial’s preregistration status. The estimated coefficients for preregistration are even slightly larger in magnitude than without these additional controls, and they are still statistically significant at the 5% level. Moreover, none of the estimated coefficients for any of these other characteristics is as large in magnitude as the coefficient of preregistration.

We conclude that the differences we established with the density discontinuity tests are indeed driven by the trial’s preregistration status and not any of these other correlated features. Moreover, of all the design characteristics commonly considered indicative of good scientific practice, preregistration appears to be the strongest signal for the credibility and integrity of reported results, as far as it can be measured with the tools at hand to detect p-hacking and selective reporting.

### Sponsor Fixed Effects

The main concern preventing a causal interpretation of the estimates of the caliper test is that there might be some unobserved trait of the researchers conducting the trials (we could call it integrity) that leads them to select both into preregistering their studies and into reporting their results completely and honestly. This selection may not be picked up by any of the observable controls we have included in the regression so far.

Attempting to control for such an unobserved trait of researchers, we consider specifications with sponsor fixed effects in columns 5 to 7 of Table 3. Comparing only preregistered and non-preregistered trials by the same sponsor, this unobserved researcher trait may cancel out as trials by the same sponsor are likely to be conducted by similar teams of researchers.^18^ Moreover, sponsor fixed effects may also account for any deliberate tampering with reported results triggered by the sponsoring party’s financial considerations.

Columns 5 and 6 repeat the first two columns’ specifications but additionally include the high-dimensional sponsor fixed effects. Column 7 controls additionally for the other design characteristics. Including all these additional controls increases the estimated difference between preregistered and non-preregistered trials to more than 12 percentage points. Standard errors increase slightly, as can be expected in such a more demanding specification with many fixed effects, but the coefficients are still statistically significant at the 10% level.

In summary, even after controlling for observables as well as possible with the available data, including sponsor fixed effects that should take care of unobserved financial considerations of sponsors and traits of responsible researchers, we find substantial and statistically significant differences between preregistered and non-preregistered trials which are indicative of preregistration being able to prevent p-hacking and selective reporting.

### Secondary Outcomes

Online Appendix C presents results from caliper tests on secondary outcomes, which confirm the results from the density discontinuity tests and do not detect any evidence of p-hacking related to these lower stake outcomes.

### Alternative Bandwidths

Table 4 shows the estimated coefficients for *preregistered* in regression equation 4 for alternative bandwidths ranging between 0.05 and 0.6. The seven columns refer to the same specifications as those in Table 3. Bandwidths lower than 0.2 deliver estimates of similar magnitude as our main specification. For the lowest bandwidths, the precision decreases, and standard errors increase naturally as the sample sizes become very small. For bandwidths larger than 0.2, the estimated effect sizes get gradually attenuated. As discussed above, the larger the bandwidth, the more likely the discontinuity estimates may be “contaminated” by differences in the underlying distributions of “true” effects.

**Table 4:** Caliper Tests with Alternative Bandwidths (Primary Outcomes, LPM)

|  | (1) | (2) | (3) | (4) | (5) | (6) | (7) |
| --- | --- | --- | --- | --- | --- | --- | --- |
| $z \in 1.96 \pm 0.05$ | -0.0874 | -0.0681 | -0.120* | -0.126 | -0.0603 | -0.101 | -0.162 |
| [$N = 254$] | (0.0633) | (0.0849) | (0.0687) | (0.0955) | (0.112) | (0.153) | (0.251) |
| $z \in 1.96 \pm 0.10$ | -0.0680 | -0.0558 | -0.0826* | -0.0871 | -0.00184 | 0.0688 | 0.0452 |
| [$N = 512$] | (0.0464) | (0.0517) | (0.0495) | (0.0566) | (0.0766) | (0.0887) | (0.114) |
| $z \in 1.96 \pm 0.15$ | -0.0895** | -0.0793* | -0.115*** | -0.117** | -0.103* | -0.0976 | -0.136* |
| [$N = 786$] | (0.0391) | (0.0440) | (0.0418) | (0.0463) | (0.0573) | (0.0706) | (0.0804) |
| $z \in 1.96 \pm 0.20$ | **-0.0862\*\*** | **-0.0740\*** | **-0.0965\*\*** | **-0.0927\*\*** | **-0.0991\*** | **-0.116\*** | **-0.121\*** |
| [$N = 1,033$] | **(0.0350)** | **(0.0396)** | **(0.0377)** | **(0.0426)** | **(0.0508)** | **(0.0614)** | **(0.0704)** |
| $z \in 1.96 \pm 0.25$ | -0.0677** | -0.0505 | -0.0754** | -0.0658* | -0.0592 | -0.0523 | -0.0538 |
| [$N = 1,327$] | (0.0316) | (0.0349) | (0.0332) | (0.0365) | (0.0442) | (0.0523) | (0.0572) |
| $z \in 1.96 \pm 0.30$ | -0.0455 | -0.0331 | -0.0554* | -0.0500 | -0.0348 | -0.0168 | -0.0191 |
| [$N = 1,585$] | (0.0287) | (0.0323) | (0.0304) | (0.0341) | (0.0402) | (0.0476) | (0.0518) |
| $z \in 1.96 \pm 0.35$ | -0.0458* | -0.0424 | -0.0511* | -0.0550* | -0.0250 | -0.0224 | -0.0146 |
| [$N = 1,826$] | (0.0274) | (0.0310) | (0.0294) | (0.0326) | (0.0373) | (0.0445) | (0.0480) |
| $z \in 1.96 \pm 0.40$ | -0.0417 | -0.0250 | -0.0471* | -0.0318 | -0.0226 | -0.00454 | 0.00475 |
| [$N = 2,102$] | (0.0256) | (0.0290) | (0.0272) | (0.0305) | (0.0344) | (0.0411) | (0.0436) |
| $z \in 1.96 \pm 0.50$ | -0.0463** | -0.0415 | -0.0510** | -0.0504* | -0.0312 | -0.0169 | -0.0199 |
| [$N = 2,645$] | (0.0235) | (0.0267) | (0.0249) | (0.0278) | (0.0305) | (0.0363) | (0.0384) |
| $z \in 1.96 \pm 0.60$ | -0.0413* | -0.0525** | -0.0372 | -0.0527* | -0.0233 | -0.0230 | -0.0232 |
| [$N = 3,135$] | (0.0229) | (0.0257) | (0.0242) | (0.0269) | (0.0287) | (0.0334) | (0.0350) |
| Controls | no | yes | no | yes | no | yes | yes |
| Start Year FE | no | yes | no | yes | no | yes | yes |
| Mesh Condition FE | no | yes | no | yes | no | yes | yes |
| Other Design Features | no | no | yes | yes | no | no | yes |
| Sponsor FE | no | no | no | no | yes | yes | yes |

Notes: The table shows the coefficient for *preregistered* in linear caliper test regressions (equation 4) on z-scores from primary outcomes in the main sample with different bandwidths in the respective rows. The columns refer to otherwise identical specifications to those in Table 3. Standard errors in parentheses are clustered at the trial level. Significance levels indicated by the stars: \* $p < 0.1$ , \*\* $p < 0.05$ , \*\*\* $p < 0.01$ .

## V Conclusion

This paper addressed whether current regulations for preregistration of pre-approval drug trials with human volunteers are sufficient to guarantee the integrity and credibility of reported results. We analyzed the distribution or results reported to the *ClinicalTrials.gov* registry from preregistered and non-preregistered trials with different tools for detecting p-hacking and selective reporting.

We do not find evidence of p-hacking in preregistered trials. In contrast, the density of z-scores from primary outcomes of non-preregistered trials exhibits patterns indicative of p-hacking or selective reporting. The difference between preregistered and non-preregistered trials regarding these indicators appears robust to conditioning on trials’ other design characteristics and sponsor fixed effects capturing unobserved researcher traits and sponsor incentives.

Preregistration at *ClinicalTrials.gov* under the current regime of regulations is indeed a reliable signal for research integrity and credibility as far as it can be evaluated with the currently available tools for detecting p-hacking and selective reporting. While we cannot provide bullet-proof evidence from exogenous or quasi-exogenous variation in the trials’ preregistration status, our results with high-dimensional sponsor fixed effects suggest that preregistration may indeed have an impact on the integrity and credibility of reported results.

As such, our results indicate that broadening preregistration requirements and stricter enforcement may further improve research credibility and integrity in the reporting of statistical evidence. However, other potential drawbacks of strict preregistration mandates, like that they lead to uninteresting and mechanical research or prevent researchers from exploring interesting ideas,^19^ should be considered by policymakers as well. A comprehensive empirical evaluation of preregistration weighing all the potential advantages and disadvantages is still an open question for future research.

## Data Availability

All data produced are available online at aact.ctti-clinicaltrials.org

## Online Appendix

### A Summary Statistics

**Table A1:**
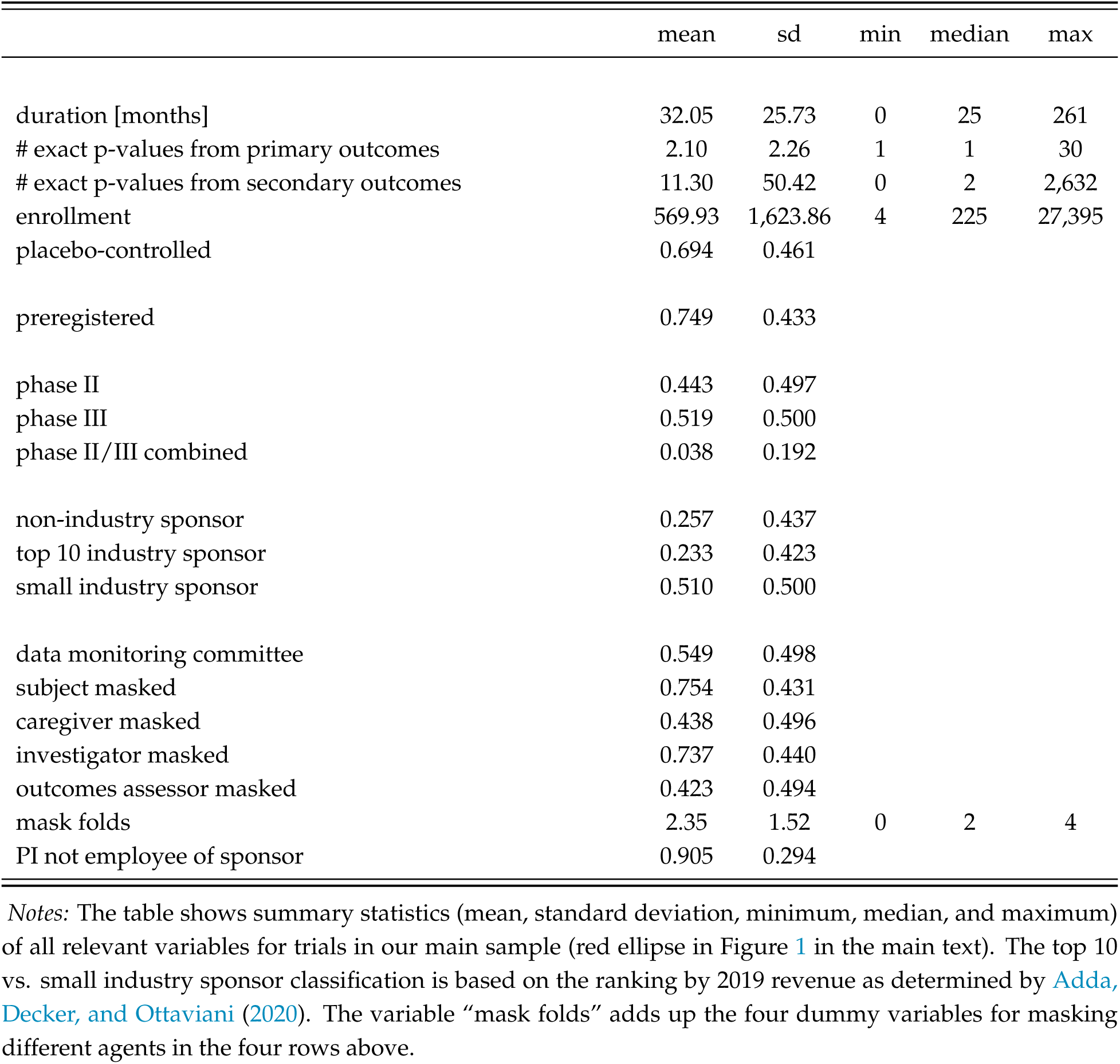
Summary Statistics for Main Sample

**Table A2:**
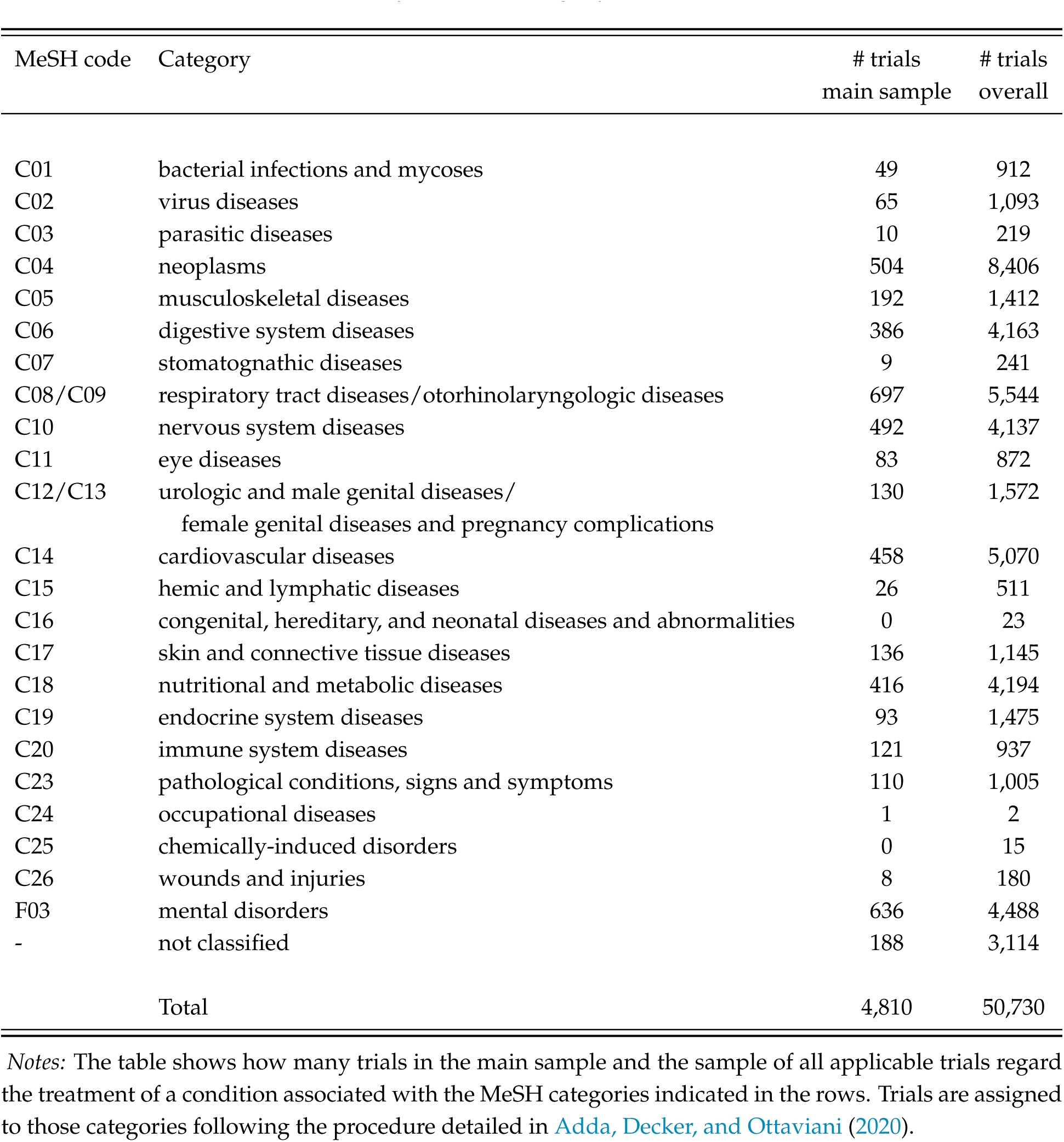
Trials by MeSH Category for Treated Condition

| MeSH code | Category | # trials<br>main sample | # trials<br>overall |
| --- | --- | --- | --- |
| C01 | bacterial infections and mycoses | 49 | 912 |
| C02 | virus diseases | 65 | 1,093 |
| C03 | parasitic diseases | 10 | 219 |
| C04 | neoplasms | 504 | 8,406 |
| C05 | musculoskeletal diseases | 192 | 1,412 |
| C06 | digestive system diseases | 386 | 4,163 |
| C07 | stomatognathic diseases | 9 | 241 |
| C08/C09 | respiratory tract diseases/otorhinolaryngologic diseases | 697 | 5,544 |
| C10 | nervous system diseases | 492 | 4,137 |
| C11 | eye diseases | 83 | 872 |
| C12/C13 | urologic and male genital diseases/<br>female genital diseases and pregnancy complications | 130 | 1,572 |
| C14 | cardiovascular diseases | 458 | 5,070 |
| C15 | hemic and lymphatic diseases | 26 | 511 |
| C16 | congenital, hereditary, and neonatal diseases and abnormalities | 0 | 23 |
| C17 | skin and connective tissue diseases | 136 | 1,145 |
| C18 | nutritional and metabolic diseases | 416 | 4,194 |
| C19 | endocrine system diseases | 93 | 1,475 |
| C20 | immune system diseases | 121 | 937 |
| C23 | pathological conditions, signs and symptoms | 110 | 1,005 |
| C24 | occupational diseases | 1 | 2 |
| C25 | chemically-induced disorders | 0 | 15 |
| C26 | wounds and injuries | 8 | 180 |
| F03 | mental disorders | 636 | 4,488 |
| - | not classified | 188 | 3,114 |
| Total |  | 4,810 | 50,730 |

*Notes:* The table shows how many trials in the main sample and the sample of all applicable trials regard the treatment of a condition associated with the MeSH categories indicated in the rows. Trials are assigned to those categories following the procedure detailed in Adda, Decker, and Ottaviani \(2020\).

**Table A3:** Comparing Characteristics of all Non-Preregistered and Preregistered Trials

|  | (1)<br>non-preregistered | (2)<br>preregistered | (3)<br>difference (2)–(1) |
| --- | --- | --- | --- |
| data monitoring committee | 0.481<br>(0.500) | 0.559<br>(0.496) | 0.078***<br>(0.004) |
| subject masked | 0.335<br>(0.472) | 0.453<br>(0.498) | 0.118***<br>(0.004) |
| caregiver masked | 0.190<br>(0.392) | 0.280<br>(0.449) | 0.090***<br>(0.003) |
| investigator masked | 0.310<br>(0.463) | 0.428<br>(0.495) | 0.117***<br>(0.004) |
| outcomes assessor masked | 0.189<br>(0.391) | 0.275<br>(0.447) | 0.086***<br>(0.003) |
| mask folds | 1.024<br>(1.502) | 1.435<br>(1.652) | 0.412***<br>(0.012) |
| PI no employee of sponsor | 0.746<br>(0.435) | 0.834<br>(0.372) | 0.088***<br>(0.006) |
| enrollment | 273.6<br>(1,695.8) | 308.6<br>(5,260.4) | 34.9<br>(32.4) |
| placebo-controlled | 0.247<br>(0.431) | 0.368<br>(0.482) | 0.121***<br>(0.003) |
| industry-sponsored | 0.405<br>(0.491) | 0.464<br>(0.499) | 0.059***<br>(0.004) |
| phase III | 0.386<br>(0.487) | 0.356<br>(0.479) | -0.030***<br>(0.004) |
| provide any result | 0.239<br>(0.426) | 0.319<br>(0.466) | 0.081***<br>(0.003) |
| provide exact p-value | 0.042<br>(0.200) | 0.076<br>(0.265) | 0.034***<br>(0.002) |
| Observations | 28,981 | 47,446 | 77,368 |

*Notes:* The first two columns show the means (and, in parentheses, standard deviations) of the variables indicated in the respective row for all applicable non-preregistered and preregistered trials in the registry (blue ellipse in Figure 1 in the main text plus ongoing trials that meet all the other criteria). Column 3 shows the differences between the first two columns with standard errors in parentheses; significance levels indicated by the stars: \* $p < 0.1$ , \*\* $p < 0.05$ , \*\*\* $p < 0.01$ . The variable “mask folds” adds up the four dummy variables in the four rows above. “Provide any result” refers to inclusion in the orange ellipse, and “provide exact p-value” refers to inclusion in the main sample, the red ellipse in Figure 1 in the main text.

### B Additional Results and Robustness Checks for Density Discontinuity Tests

This section presents additional results from density discontinuity tests and robustness checks of the main results presented in the main text.

#### Extent of Registration Delay

Our main analysis uses a binary variable to indicate if a study has been preregistered (registration date to the registry before or equal to the start date of the trial) or non-preregistered (registration date to the registry after the start date of the trial). Figure B1 looks at the distribution of registration delays defined as registration date minus start date. This means preregistered trials have a registration delay *≤* 0, and non-preregistered trials have a positive registration delay.

Panel A shows a histogram of the registration delay in days of all trials in our sample. Most trials register in the weeks just preceding the start date, with the mode of the distribution at -7 days. Most non-preregistered trials are only delayed by a few weeks, while the distribution has a long tail of delays of several hundred days, which in the graph is truncated at 500 days corresponding only to the 94th percentile of the distribution.

Panel B zooms into the window of delays between 1 and 50 days. There is no bunching of registrations before the 21-day threshold to comply with the FDA registration requirements.

Panel C shows the registration delay relative to the trial duration defined as primary completion date minus start date (i.e., a relative delay >1 means that the trial was only registered after completion). The distribution is very similar to the distribution of absolute delays in panel A.

The distribution of registration delays begs the question if trials that register only with a slight delay appear less p-hacked than trials with a more substantial delay. If a trial is registered with a delay of a few days only, this might be due to oversight rather than intention. The investigators most likely do not have information yet based on which they could modify the trial protocol or outcome measures to inflate statistical significance. More substantial delays may be more likely to be intentional, and investigators may already have more information from interim analyses based on which they could modify the trial protocol or outcome measures to achieve a higher level of statistical significance before the first registration. The larger the registration delay, the more opportunity there is for p-hacking that cannot be caught by comparing initial registration with the final reports.

In Figure B2, we repeat the density discontinuity tests for primary outcomes at *z* = 1.96, splitting the sample of non-preregistered trials by their relative registration delay. The cutoff of 10% relative registration delay (0.1 in the distribution plotted in panel C of Figure B1), splits the sample non-preregistered trials roughly in half, maximizing the power for the demanding discontinuity test in both groups.

It is striking that the discontinuity appears to be driven almost entirely by the trials with a registration delay of more than 10% of the trial duration (panel B). Especially the spike in the bin right above 1.96, which may be seen as evidence for inflation of results to push them just over the threshold, is extremely pronounced for the strongly delayed trials and completely absent for the slightly delayed trials (panel A).

Even though with the halved sample size the density discontinuity test is underpowered to detect a statistically significant break for either of the two groups, we see the pattern that the discontinuity grows with the opportunity to p-hack as indicative evidence that the discontinuity is indeed driven by some sort of manipulation.

#### Other Significance Thresholds

While 5% is the most prominent and salient threshold for the evaluation of statistical significance, some studies could also p-hack to clear a different threshold. Other commonly used thresholds to claim statistical significance or assign ’stars’ in result tables include 10%, 1%, and 0.1%.

Figure B3 shows density discontinuity tests for the distribution of z-scores from tests on primary outcomes at the threshold *z* = 1.64, corresponding to statistical significance at the 10% level. Neither the test detects a significant discontinuity for non-preregistered trials (panel A) nor for preregistered trials (panel B).

Figure B4 shows density discontinuity tests for the distribution of z-scores from tests on primary outcomes at the threshold *z* = 2.58, corresponding to statistical significance at the 1% level. The density from non-preregistered trials (panel A) displays a substantial downward jump at this threshold, even though not statistically significant. This pattern is mainly driven by the large mass of trials with z-scores just above 1.96, pushing the density estimate upwards to the left of the *z* = 2.58 threshold. The density from preregistered trials (panel B) appears smooth at this threshold.

Figure B5 shows density discontinuity tests for the distribution of z-scores from tests on primary outcomes at the threshold *z* = 3.29, corresponding to statistical significance at the 0.1% level. Both the density from non-preregistered trials (panel A) and the density from preregistered trials (panel) B exhibit a statistically significant downward jump at this threshold. We attribute this pattern to the fact that we only consider exactly reported p-values, and most p-values smaller than 0.001 (corresponding to z-scores to the right of the tested threshold) are not reported exactly but only as *p <* 0.001 and *p <* 0.0001. These p-values are not included in the sample for Figure B5, and the density estimate is based on only a small part of the actual mass of z-scores to the right of the threshold (see the discussion in footnote 8 of the main text). Therefore, the downward discontinuity is not surprising.

In summary, density discontinuity tests do not detect evidence for p-hacking or selective reporting at any of the considered alternative significance thresholds, neither for non-preregistered nor preregistered trials. However, the tests at the 0.1% threshold should be interpreted with a grain of salt, as we only have very little information about the shape of the very right tail of the z-densities.

#### Other Trial Characteristics

As discussed in the main text, there are design elements of trials other than preregistration, which are usually considered signs of research integrity, such as blinding or the presence of a data monitoring committee. These elements are correlated with the trials’ preregistration status (see Table 1 in the main text). The density discontinuity tests do not allow for an evaluation conditional on other covariates but force us to do discrete sample splits if we want to assess the impact of a specific variable on the discontinuity.

Table B1 shows the log-difference measure Δ_1.96_ and, in parentheses, p-values of discontinuity tests at *z* = 1.96 of z-scores pertaining to primary outcomes for binary sample splits based on these other design elements. All variables are coded so that ’yes’ corresponds to the superior designs.

Neither of these other design elements appears to have an impact on the discontinuity indicative of p-hacking which is as stark as the impact of the lack of preregistration. If anything, the trials with superior design features tend to exhibit larger jumps. As noted by Adda, Decker, and Ottaviani (2020), trials with superior design features are often those with a higher prior of success ex-ante and, therefore, more significant results ex-post. The density discontinuity tests might pick up partially this difference in the underlying distribution of true effects.

To control for the impact of all of these features simultaneously, we apply caliper tests (Section IV of the main text).

#### Robustness Check: Transformation to One-Sided Test Statistics

To rule out that the discontinuity we find for z-scores from non-preregistered trials at the 5% significance threshold is not driven by the particular transformation we apply to the reported p-values, we consider an alternative transformation by supposing that the p-values do not originate from a two-sided but a one-sided Z test. This alternative transformation corresponds to the one-to-one mapping

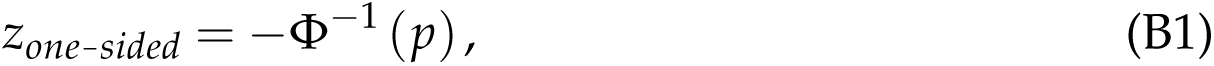

where *−***Φ***^−^*^1^ is the inverse of the standard normal cumulative distribution function.

For a one-sided Z test, the 5% significance threshold is at *z_one-sided_* = 1.64. Figure B6 shows density discontinuity tests for the transformed one-sided test statistics from primary outcomes of non-preregistered trials (panel A) and preregistered trials (panel B) at this threshold. The discontinuity estimates are similar in size and statistical significance level to those from the transformation to z-scores from a two-sided test (Figure 4 in the main text).

#### Robustness Check: De-Rounding P-Values

A potential concern in the literature that analyzes the distribution of z-scores is that rounding of parameter estimates, standard errors, test statistics, or p-values could influence the estimation of densities and discontinuity tests, as rounding might lead to the accumulation of mass at specific values. For instance, papers that construct z-scores directly from parameter estimates and standard errors in published articles are concerned about rounding leading to many z-scores of exactly 2, right above the significance threshold of 1.96, which might actually be below 1.96 if reported more precisely (Brodeur et al., 2016; Kranz and Pütz, 2022). To test for the robustness of estimates with respect to such rounding issues, de-rounding procedures have been proposed.

As we do not construct our z-score from parameter estimates and standard errors but ’backward’ from p-values, misclassifying a p-value as statistically significant based on rounding appears less concerning. Rounding a p-value greater than 0.05 to 0.05 or less and reporting it explicitly as a statistically significant p-value of less or equal to 0.05 can be seen as a form of p-hacking in itself. Therefore, such a practice would not undermine the validity of our tests for manipulation, but it *should* be picked up as evidence for manipulation. Moreover, note that our sample of z-scores constructed from p-values contains no z-score of exactly 2.

However, rounding of p-values could still affect our density estimation to some extent. To test for the robustness of our results in this regard, we apply a de-rounding procedure similar to the one proposed by Brodeur et al. (2016) directly to the reported p-values (instead of de-rounding parameter estimates and standard errors as in their original application). To construct a sample of de-rounded p-values, we replace each p-value with a random draw from a uniform distribution over the interval that would be rounded to the reported value, as long as this replacement would not turn a significant result (*p <* 0.05) into an insignificant result (*p >* 0.05). We make this latter restriction because, as discussed above, rounding an insignificant p-value to a significant one could by itself be seen as a form of p-hacking, and, therefore, should be picked up by a test for manipulation. For example, a p-value reported as 0.06 will be replaced by a random draw from the interval [0.55, 0.65), and a p-value reported as 0.032 will be replaced by a random draw from the interval [0.0315, 0.0325). We then transform these de-rounded p-values into corresponding z-scores according to equation 1 in the main text.

Figure B7 repeats the density discontinuity tests at *z* = 1.96 for primary outcomes with the z-scores obtained from the de-rounded p-values. The size and precision of the estimated discontinuity for non-preregistered trials (panel A) are attenuated only slightly compared to the results in Figure 4 in the main text.

#### Alternative Tests to Detect p-Hacking Proposed by Elliott, Kudrin, and Wüthrich (2022)

Table B2 shows p-values for the battery of tests to detect p-hacking proposed by Elliott, Kudrin, and Wüthrich (2022). These tests are based on several properties that the distribution of p-values should exhibit under general conditions in the absence of p-hacking, such as continuity, non-increasingness, and *K*-monotonicity.

- ‘Fisher’ refers to a Fisher’s test of the null hypothesis that the density of p-values is non-increasing (Simonsohn, Nelson, and Simmons, 2014).
- ‘Binomial’ refers to a Binomial test of the non-increasingness of the density of p-values on the interval [0.04, 0.05], which compares the number of p-values in the two subintervals [0.04, 0.045] and (0.045, 0.05].
- ‘CS1’ refers to a histogram-based test for the non-increasingness of the density of p-values, which is implemented using the conditional chi-squared test of Cox and Shi (2023).
- ‘CS2B’ refers to a histogram-based test for 2-monotonicity and bounds on the density of p-values and the first two derivatives, which is implemented using the conditional chi-squared test of Cox and Shi (2023).
- ‘Discontinuity’ refers to the density discontinuity test by Cattaneo, Jansson, and Ma (2020) applied to the density of p-values at the threshold 0.05.
- ‘LCM’ refers to a Least Concave Majorant test of the null hypothesis that the cumulative distribution function of p-values is convex.

Note that all these tests operate on the distribution of p-values, unlike the tests presented in the main text, which are applied to the distribution of z-scores. Moreover, we follow Elliott, Kudrin, and Wüthrich (2022) and truncate the distribution of p-values above 0.15 for all tests.

The first two columns show the results for primary outcomes of non-preregistered and preregistered trials, respectively. The third column shows average results from tests performed on 100 subsamples of p-values from preregistered trials of size equal to the number of observations from non-preregistered trials. This downsampling procedure allows for a comparison of p-values between non-preregistered and preregistered trials that is not driven by the different sample sizes.

For non-preregistered trials (column 1) and the average subsample of preregistered trials (column 3), all tests other than ’CS2B’ do not reject the null hypothesis (no indication of p-hacking) at a 95% confidence level. In the full sample of preregistered trials (column 2), additionally, the ’CS1’ test rejects the null. However, due to the three times as large sample size, we naturally expect smaller p-values than for the other samples.

For our main analysis, we rely on discontinuity tests of the density of z-scores and caliper tests. Compared to the tests presented in Table B2, discontinuity tests of the zdensity have the advantage that they do not require truncating the distribution of results ex-ante but potentially exploit information of the entire distribution in a data-driven manner. Moreover, the estimator, which relies on a locally quadratic and cubic approximation of the density, can identify discontinuities more reliably in the z-density, which is close to linear at the significance threshold in the absence of manipulation, than in the p-density, which is highly convex around the threshold. In consequence, discontinuity tests on the z-density yield results that are most aligned with an intuitive visual evaluation of the histograms of the raw data. Caliper tests have the advantage over the tests presented in Table B2 that they allow to condition on other covariates.

**Figure B1:**
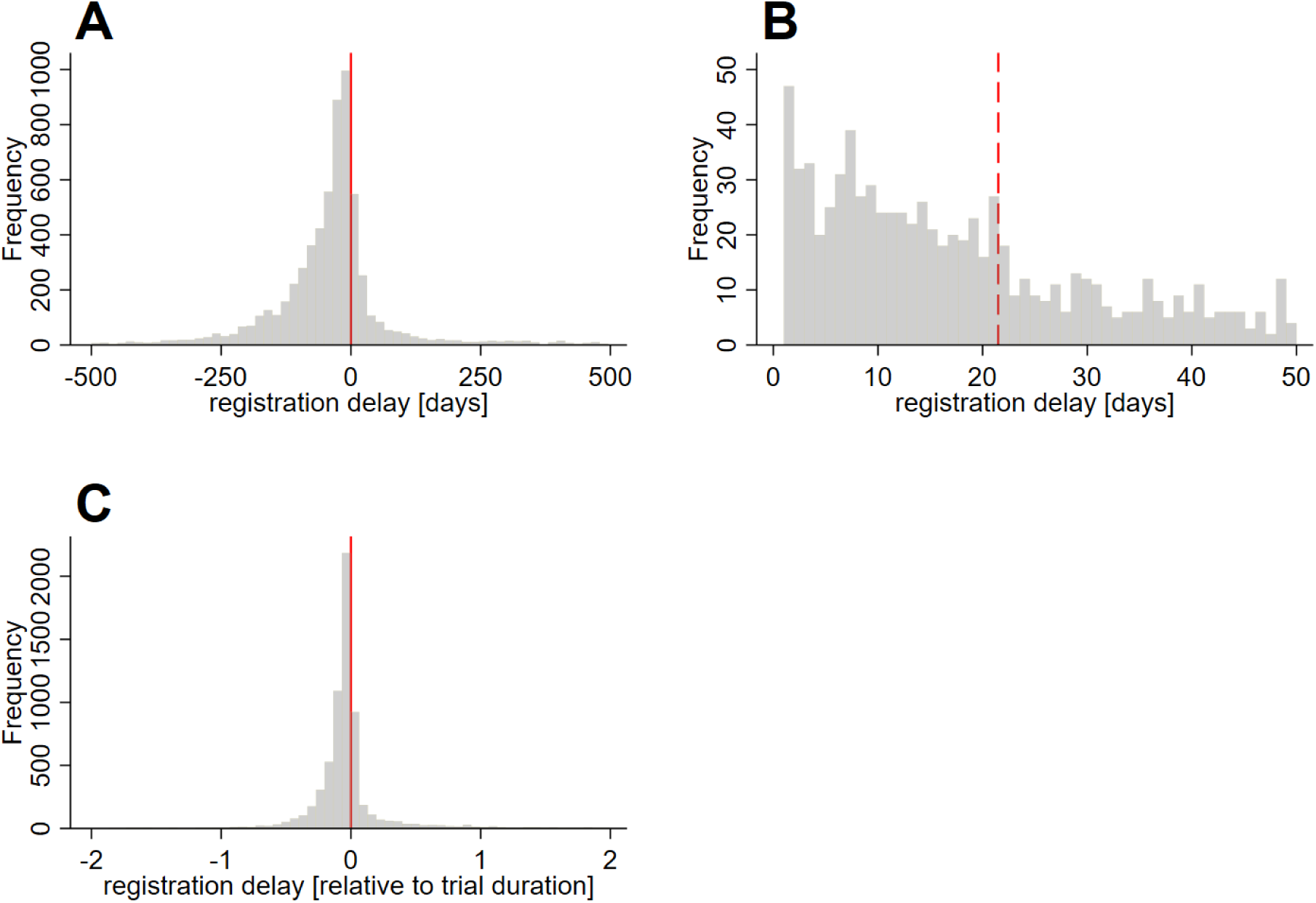
Registration Delay *Notes:* The graphs show histograms for the distribution of registration delays, defined as start date minus registration date, such that preregistered trials have a negative registration delay and non-preregistered trials have a positive registration delay. Panels A and B show absolute registration delays in days. Panel C shows the relative registration delay with respect to the trial duration. The solid vertical red lines in panels A and C mark the preregistration threshold of zero registration delay. The dashed vertical red line in panel B marks a registration delay of 21 days, which is the threshold to comply with FDA regulations (see discussion in Section II of the main text).

**Figure B2:**
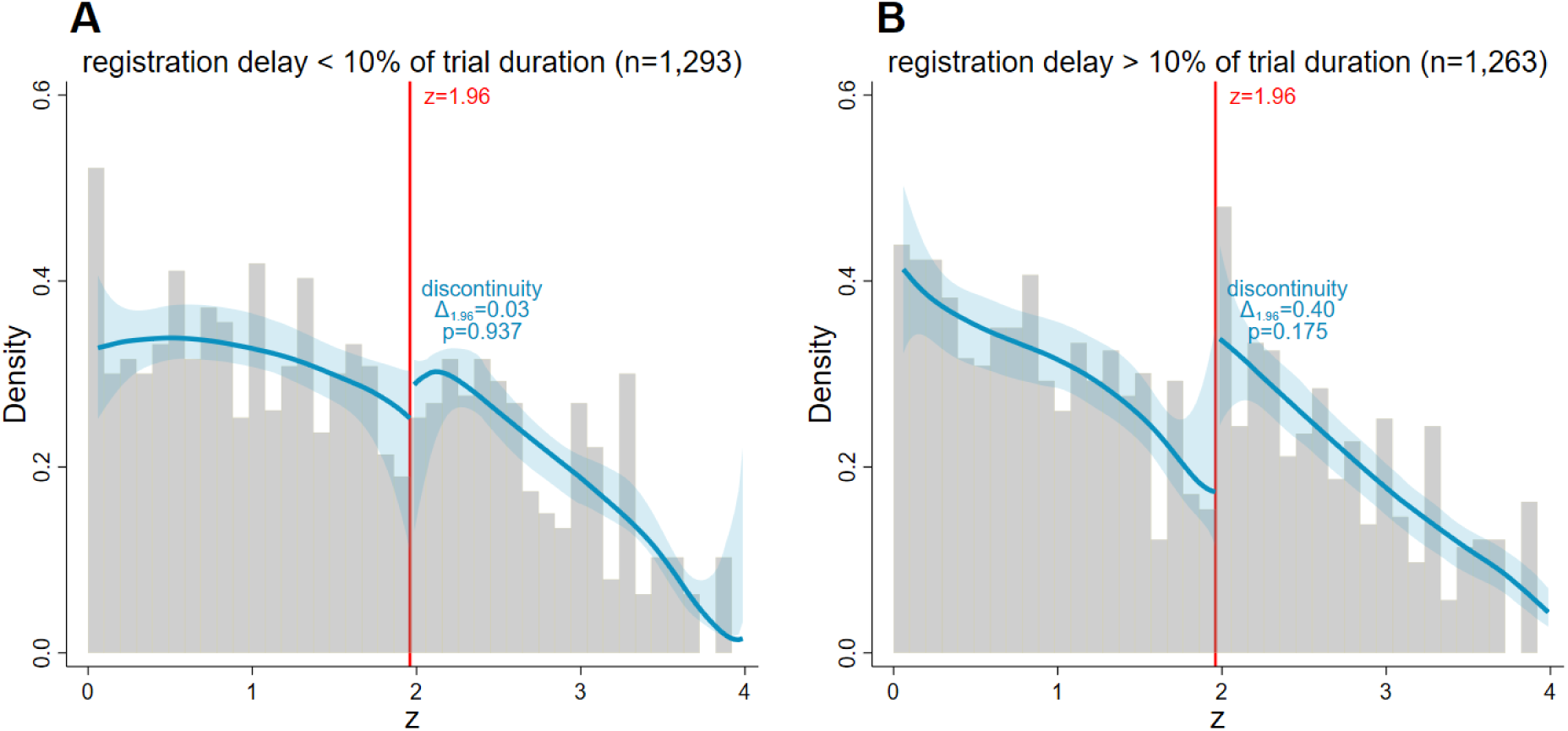
Registration Delay: Density Discontinuity Tests at *z* = 1.96 (Primary Outcomes) *Notes:* The graphs show histograms (grey) and density estimates (blue) for the distribution of z-scores from tests on primary outcomes of trials registered with a delay of 0-10% of the trial duration (panel A) and more than 10% (panel B). The shaded blue areas are 95% confidence bands for the density estimates, and the vertical red lines at 1.96 correspond to the threshold for statistical significance at the 0.05 level. The local polynomial density estimators proposed by Cattaneo, Jansson, and Ma (2020) are used. We allow for a discontinuity at 1.96 and present the log-difference measure Δ_1.96_ as defined in equation 3 in the main text and the p-values of the discontinuity test (equation 2 in the main text). Note that potential discrepancies between Δ_1.96_ and the differences that can be read off the plotted densities are due to the bias correction in the testing procedure.

**Figure B3:**
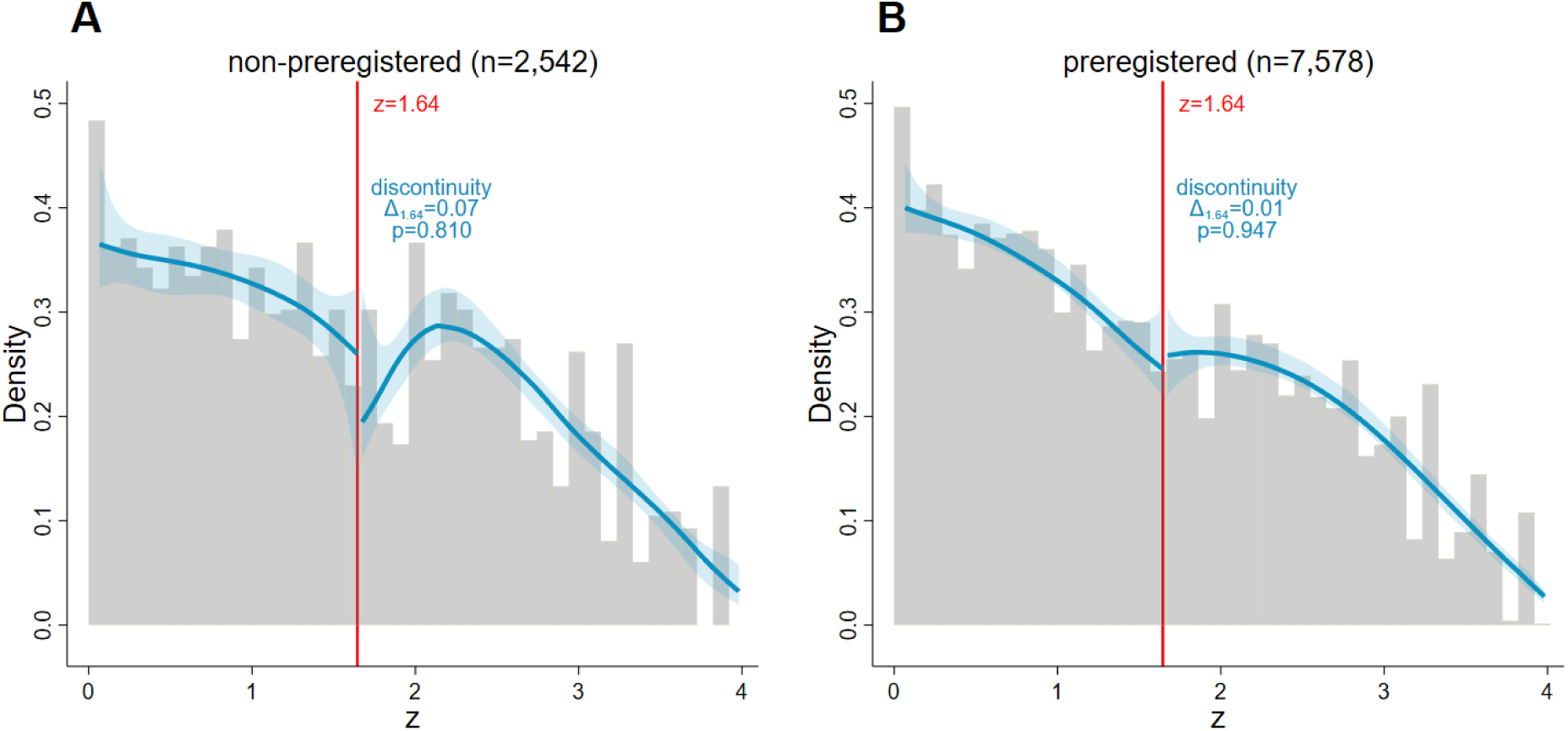
Density Discontinuity Tests at *z* = 1.64 (Primary Outcomes) *Notes:* The graphs show histograms (grey) and density estimates (blue) for the distribution of z-scores from tests on primary outcomes of non-preregistered (panel A) and preregistered trials (panel B). The shaded blue areas are 95% confidence bands for the density estimates, and the vertical red lines at 1.64 correspond to the threshold for statistical significance at the 0.1 level. The local polynomial density estimators proposed by Cattaneo, Jansson, and Ma (2020) are used. We allow for a discontinuity at 1.64 and present the log-difference measure Δ_1.64_ as defined in equation 3 in the main text and the p-values of the discontinuity test (equation 2 in the main text). Note that potential discrepancies between Δ_1.64_ and the differences that can be read off the plotted densities are due to the bias correction in the testing procedure. The density estimates in panel A rely on local cubic approximation instead of local quadratic approximation, which is used in all the other estimates, to allow the necessary flexibility to appropriately fit the sharp hump around *z* = 2.

**Figure B4:**
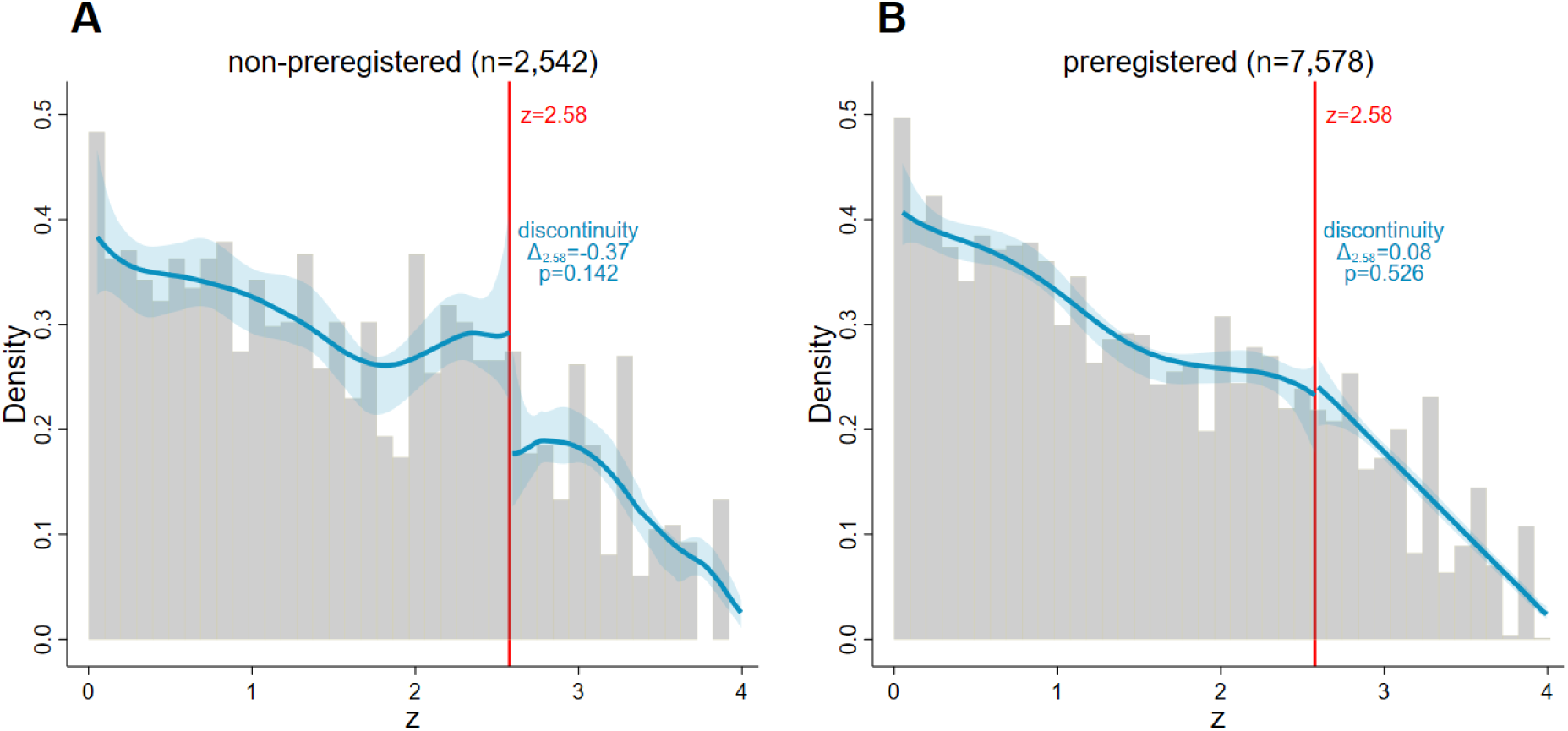
Density Discontinuity Tests at *z* = 2.58 (Primary Outcomes) *Notes:* The graphs show histograms (grey) and density estimates (blue) for the distribution of z-scores from tests on primary outcomes of non-preregistered (panel A) and preregistered trials (panel B). The shaded blue areas are 95% confidence bands for the density estimates, and the vertical red lines at 2.58 correspond to the threshold for statistical significance at the 0.01 level. The local polynomial density estimators proposed by Cattaneo, Jansson, and Ma (2020) are used. We allow for a discontinuity at 2.58 and present the log-difference measure Δ_2.58_ as defined in equation 3 in the main text and the p-values of the discontinuity test (equation 2 in the main text). Note that potential discrepancies between Δ_2.58_ and the differences that can be read off the plotted densities are due to the bias correction in the testing procedure.

**Figure B5:**
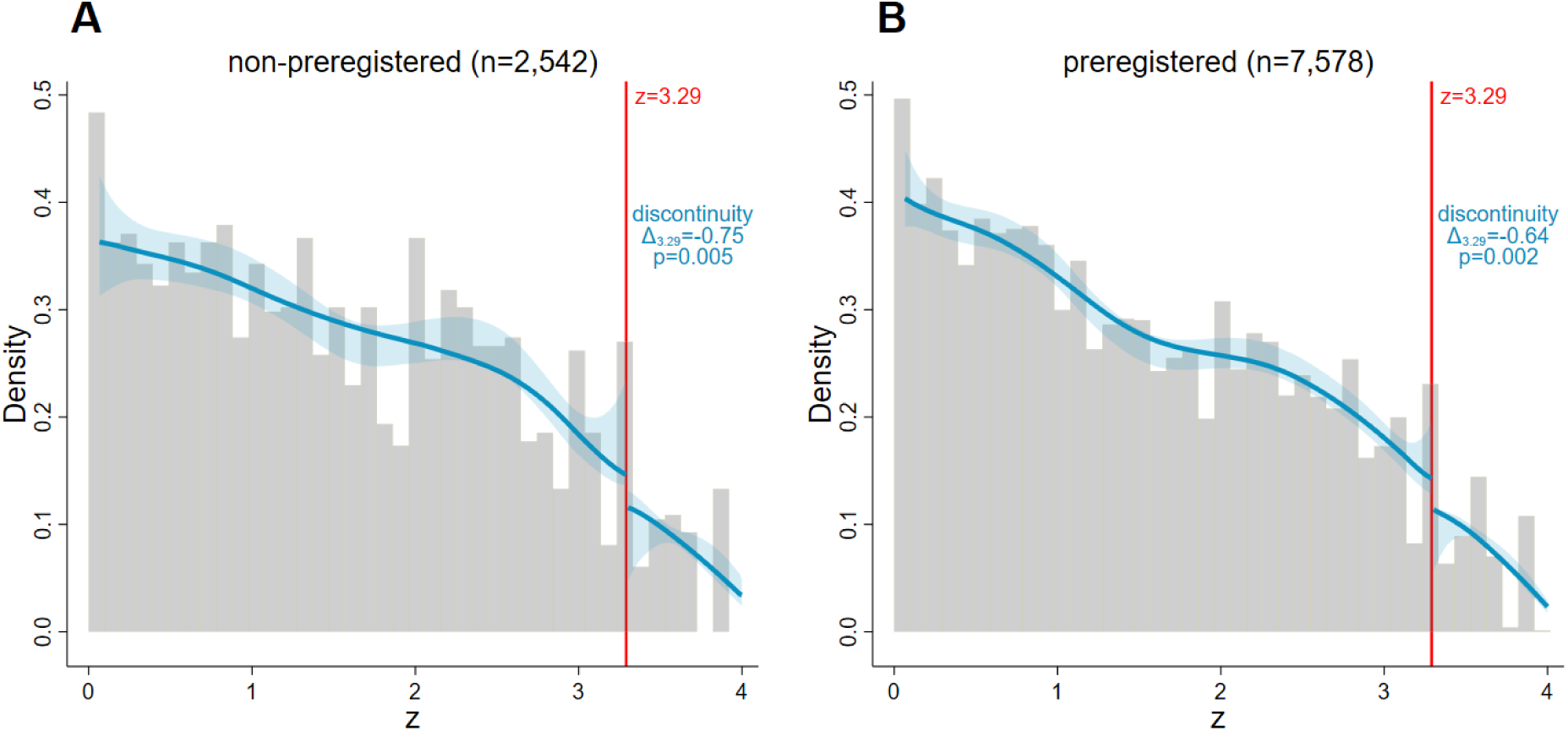
Density Discontinuity Tests at *z* = 3.29 (Primary Outcomes) *Notes:* The graphs show histograms (grey) and density estimates (blue) for the distribution of z-scores from tests on primary outcomes of non-preregistered (panel A) and preregistered trials (panel B). The shaded blue areas are 95% confidence bands for the density estimates, and the vertical red lines at 3.29 correspond to the threshold for statistical significance at the 0.001 level. The local polynomial density estimators proposed by Cattaneo, Jansson, and Ma (2020) are used. We allow for a discontinuity at 3.29 and present the log-difference measure Δ_3.29_ as defined in equation 3 in the main text and the p-values of the discontinuity test (equation 2 in the main text). Note that potential discrepancies between Δ_3.29_ and the differences that can be read off the plotted densities are due to the bias correction in the testing procedure.

**Figure B6:**
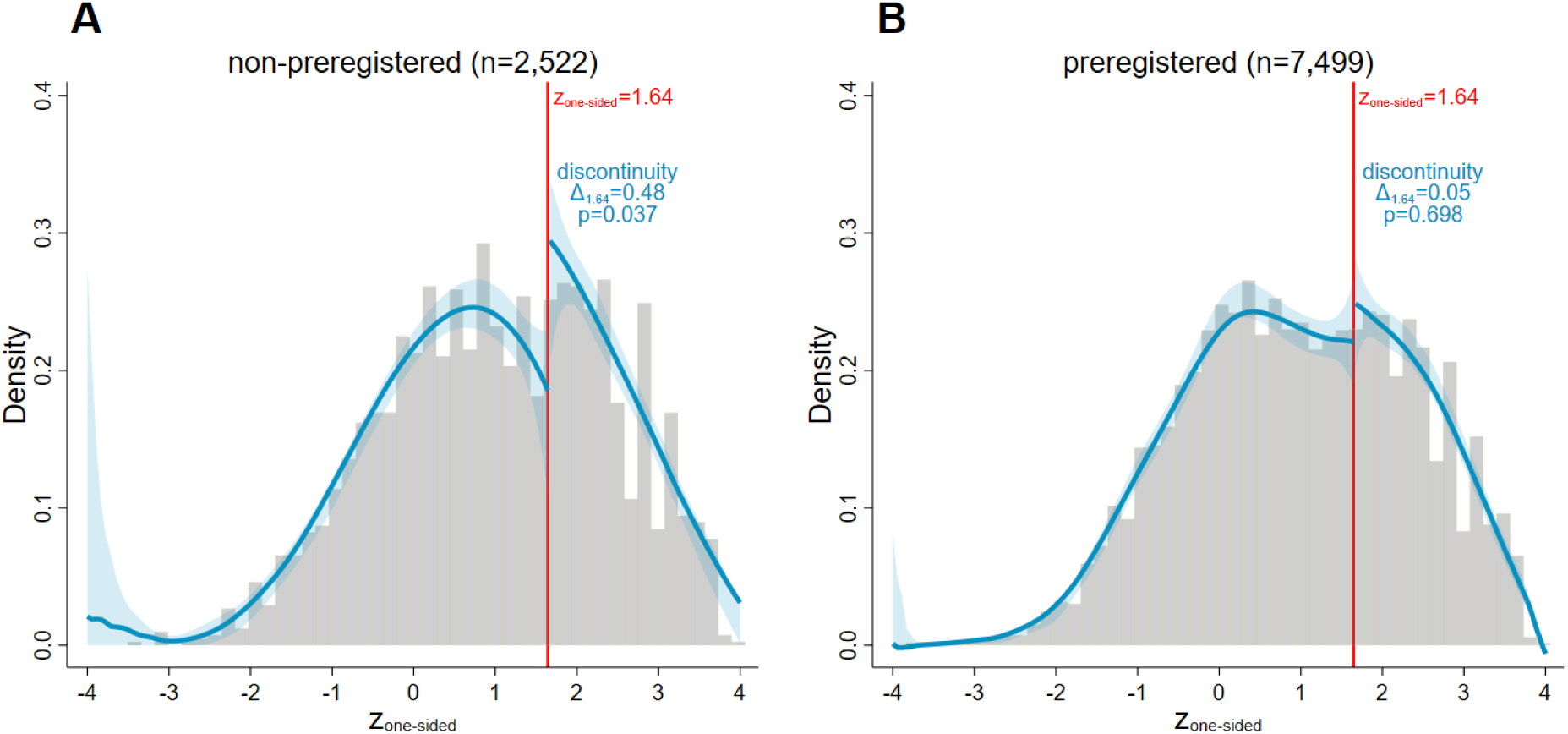
Density Discontinuity Tests at *z_one-sided_* = 1.64 (Primary Outcomes) *Notes:* The graphs show histograms (grey) and density estimates (blue) for the distribution of transformed one-sided z-scores from tests on primary outcomes of non-preregistered (panel A) and preregistered trials (panel B). The shaded blue areas are 95% confidence bands for the density estimates, and the vertical red lines at 1.64 correspond to the threshold for statistical significance at the 0.05 level of a one-sided test. The local polynomial density estimators proposed by Cattaneo, Jansson, and Ma (2020) are used. We allow for a discontinuity at 1.64 and present the log-difference measure Δ_1.64_ as defined in equation 3 in the main text and the p-values of the discontinuity test (equation 2 in the main text). Note that potential discrepancies between Δ_1.64_ and the differences that can be read off the plotted densities are due to the bias correction in the testing procedure.

**Figure B7:**
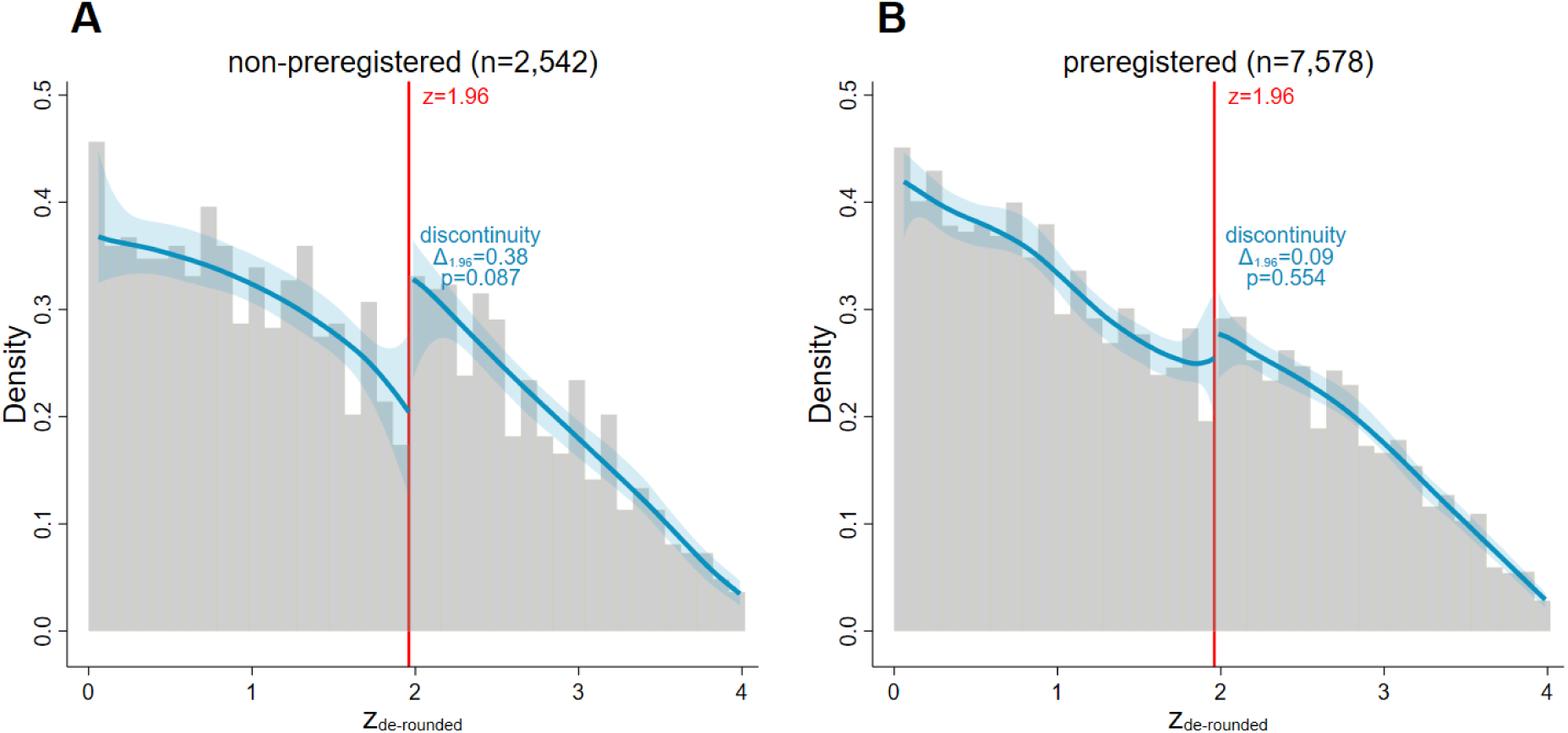
De-Rounding: Density Discontinuity Tests at *z_de-rounded_* = 1.96 (Primary Outcomes) *Notes:* The graphs show histograms (grey) and density estimates (blue) for the distribution of z-scores, calculated from p-values de-rounded as detailed in the text, from tests on primary outcomes of non-preregistered (panel A) and preregistered trials (panel B). The shaded blue areas are 95% confidence bands for the density estimates, and the vertical red lines at 1.96 correspond to the threshold for statistical significance at the 0.05 level. The local polynomial density estimators proposed by Cattaneo, Jansson, and Ma (2020) are used. We allow for a discontinuity at 1.96 and present the log-difference measure Δ_1.96_ as defined in equation 3 in the main text and the p-values of the discontinuity test (equation 2 in the main text). Note that potential discrepancies between Δ_1.96_ and the differences that can be read off the plotted densities are due to the bias correction in the testing procedure.

**Table B1:**
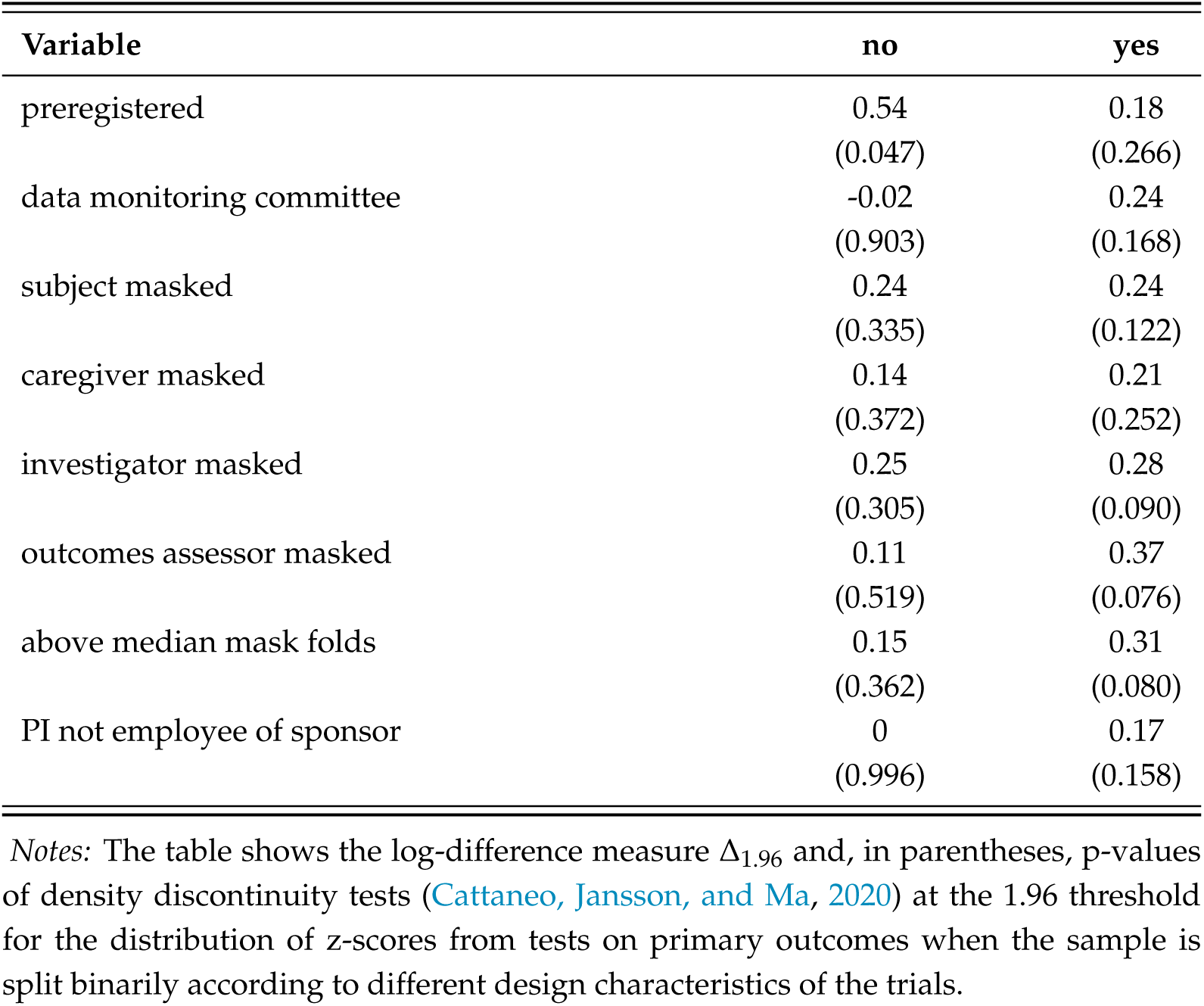
Other Trial Characteristics: Δ_1.96_ and P-Values of Density Discontinuity Tests

| Variable | no | yes |
| --- | --- | --- |
| preregistered | 0.54<br>(0.047) | 0.18<br>(0.266) |
| data monitoring committee | -0.02<br>(0.903) | 0.24<br>(0.168) |
| subject masked | 0.24<br>(0.335) | 0.24<br>(0.122) |
| caregiver masked | 0.14<br>(0.372) | 0.21<br>(0.252) |
| investigator masked | 0.25<br>(0.305) | 0.28<br>(0.090) |
| outcomes assessor masked | 0.11<br>(0.519) | 0.37<br>(0.076) |
| above median mask folds | 0.15<br>(0.362) | 0.31<br>(0.080) |
| PI not employee of sponsor | 0<br>(0.996) | 0.17<br>(0.158) |

*Notes:* The table shows the log-difference measure $\Delta_{1.96}$ and, in parentheses, p-values of density discontinuity tests (Cattaneo, Jansson, and Ma, 2020) at the 1.96 threshold for the distribution of z-scores from tests on primary outcomes when the sample is split binarily according to different design characteristics of the trials.

**Table B2:** Tests for the Detection of p-Hacking Proposed by Elliott, Kudrin, and Wüthrich (2022)

|  | (1)<br>non-preregistered | (2)<br>preregistered | (3)<br>preregistered<br>(downsampled) |
| --- | --- | --- | --- |
| Fisher | 1.000 | 1.000 | 1.000 |
| Binomial | 0.336 | 0.275 | 0.399 |
| CS1 | 0.129 | 0.000 | 0.059 |
| CS2B | 0.001 | 0.000 | 0.023 |
| Discontinuity | 0.804 | 0.054 | 0.216 |
| LCM | 0.536 | 0.173 | 0.811 |
| # p-values in $[0.04, 0.05]$ | 89 | 226 | 76.8 |
| # p-values in $[0.00, 0.15]$ | 1,280 | 3,657 | 1,229.2 |
| # p-values | 2,542 | 7,578 | 2,542 |

*Notes:* The table shows p-values for the different tests of p-hacking for the samples of primary outcomes. ‘Discontinuity’ refers to a density discontinuity test (Cattaneo, Jansson, and Ma, 2020) of the density of p-values at the cutoff of 0.05. ‘Fisher’ refers to Fisher’s test. ‘Binomial’ refers to a binomial test on the interval $[0.04, 0.05]$ . ‘CS1’ refers to a conditional chi-squared test (Cox and Shi, 2023) for non-increasingness. ‘CS2B’ refers to a conditional chi-squared test (Cox and Shi, 2023) for 2-monotonicity and bounds (Elliott, Kudrin, and Wüthrich, 2022). ‘LCM’ refers to the Least Concave Majorant test. We follow Elliott, Kudrin, and Wüthrich \(2022\) and truncate the distribution of p-values above 0.15 for all tests. The third column shows average results from tests performed on 100 random subsamples of preregistered trials of size equal to the number of observations from non-preregistered trials.

### C Additional Results and Robustness Checks for Caliper Tests

This section presents additional results from caliper tests and robustness checks of the main results presented in the main text.

#### Secondary Outcomes

Table C1 shows the results of caliper tests for z-scores from secondary outcomes for the same specifications as presented in Table 3 in the main text for primary outcomes. As the density discontinuity tests, the caliper tests do not detect major differences between preregistered and non-preregistered trials related to these lower-stake auxiliary outcomes. If anything, when introducing the control variables, preregistered trials tend to have slightly more significant z-scores in the window under consideration than non-preregistered trials. However, the magnitude of the differences is much smaller than for primary outcomes.

#### Robustness Check: Alternative Model Specifications

Tables C2 and C3 show the marginal effects of probit and logit regressions for caliper tests, respectively. The specifications include the same variables as those presented in the first four columns of Table 3 in the main text; those are the specifications without the high-dimensional sponsor fixed effects. The estimated coefficients of interest are nearly identical to those resulting from our main specification of a linear probability model.

**Table C1:** Caliper Tests for *z ∈*1.96*±*0.2 (Secondary Outcomes, LPM)

|  | (1) | (2) | (3) | (4) | (5) | (6) | (7) |
| --- | --- | --- | --- | --- | --- | --- | --- |
| preregistered | 0.00679<br>(0.0159) | 0.0225<br>(0.0185) | 0.0233<br>(0.0172) | 0.0395**<br>(0.0197) | 0.0213<br>(0.0198) | 0.0420*<br>(0.0233) | 0.0623**<br>(0.0250) |
| data monitoring committee |  |  | -0.0145<br>(0.0166) | -0.0223<br>(0.0185) |  |  | -0.0230<br>(0.0233) |
| subject masked |  |  | -0.104*<br>(0.0611) | -0.0748<br>(0.0686) |  |  | -0.0542<br>(0.119) |
| caregiver masked |  |  | 0.0210<br>(0.0176) | 0.0277<br>(0.0199) |  |  | 0.0314<br>(0.0273) |
| investigator masked |  |  | 0.0459<br>(0.0569) | 0.0727<br>(0.0554) |  |  | 0.0500<br>(0.108) |
| outcomes assessor masked |  |  | -0.0114<br>(0.0172) | -0.0112<br>(0.0196) |  |  | -0.00896<br>(0.0252) |
| PI not employee of sponsor |  |  | 0.00647<br>(0.0505) | 0.0330<br>(0.0573) |  |  | -0.00417<br>(0.0910) |
| Non-Prereg. Sig. Rate | 0.481 | 0.481 | 0.471 | 0.471 | 0.482 | 0.482 | 0.469 |
| Observations | 5,149 | 5,149 | 4,602 | 4,602 | 5,004 | 5,004 | 4,462 |
| No. of trials | 1,458 | 1,458 | 1,266 | 1,266 | 1,313 | 1,313 | 1,126 |
| R-squared | 0.000 | 0.014 | 0.002 | 0.018 | 0.057 | 0.067 | 0.074 |
| Controls | no | yes | no | yes | no | yes | yes |
| Start Year FE | no | yes | no | yes | no | yes | yes |
| MeSH Condition FE | no | yes | no | yes | no | yes | yes |
| Sponsor FE | no | no | no | no | yes (243) | yes (243) | yes (227) |
| Window | 1.96±0.2 | 1.96±0.2 | 1.96±0.2 | 1.96±0.2 | 1.96±0.2 | 1.96±0.2 | 1.96±0.2 |

*Notes:* The table shows results from the linear caliper test regression (equation 4 in the main text) on z-scores from secondary outcomes in the main sample. The seven specifications are identical to those in Table 3 in the main text. “Controls” include dummies for research phases (phase II vs. phase III vs. phase II/III combined), a dummy for a placebo comparator, the square root of overall enrollment as a proxy for the power of the tests, and in the specifications without sponsor fixed effects dummies for sponsor groups (non-industry, small industry, and top 10 industry based on 2019 revenue as defined by Adda, Decker, and Ottaviani \(2020\)). “MeSH Condition FE” refers to fixed effects for groups of treated definitions based on MeSH classification (see Table A2; categories with too few observations are grouped together as omitted category). Reduced sample sizes result from excluding trials with missing information on data monitoring committees in columns 3, 4, and 7 and excluding singleton observations within sponsor nests in columns 5, 6, and 7. Standard errors in parentheses are clustered at the trial level. Significance levels indicated by the stars: \* $p < 0.1$ , \*\* $p < 0.05$ , \*\*\* $p < 0.01$ .

**Table C2:** Caliper Tests for *z ∈*1.96*±*0.2 (Primary Outcomes, Marginal Effects from Probit Model without Sponsor FE)

|  | (1) | (2) | (3) | (4) |
| --- | --- | --- | --- | --- |
| preregistered | -0.0869**<br>(0.0356) | -0.0737*<br>(0.0392) | -0.0979**<br>(0.0383) | -0.0918**<br>(0.0417) |
| data monitoring committee |  |  | 0.0254<br>(0.0330) | 0.00283<br>(0.0352) |
| subject masked |  |  | 0.0108<br>(0.0880) | -0.0155<br>(0.0893) |
| caregiver masked |  |  | 0.00752<br>(0.0440) | 0.00449<br>(0.0428) |
| investigator masked |  |  | -0.0796<br>(0.0848) | -0.0449<br>(0.0806) |
| outcomes assessor masked |  |  | 0.0418<br>(0.0424) | 0.0620<br>(0.0413) |
| PI not employee of sponsor |  |  | -0.0185<br>(0.0611) | 0.0766<br>(0.0729) |
| Non-Prereg. Sig. Rate | 0.633 | 0.620 | 0.638 | 0.623 |
| Observations | 1,033 | 1,024 | 912 | 903 |
| No. of trials | 850 | 842 | 747 | 738 |
| Controls | no | yes | no | yes |
| Start Year FE | no | yes | no | yes |
| MeSH Condition FE | no | yes | no | yes |
| Window | 1.96±0.2 | 1.96±0.2 | 1.96±0.2 | 1.96±0.2 |

**Table C3:**
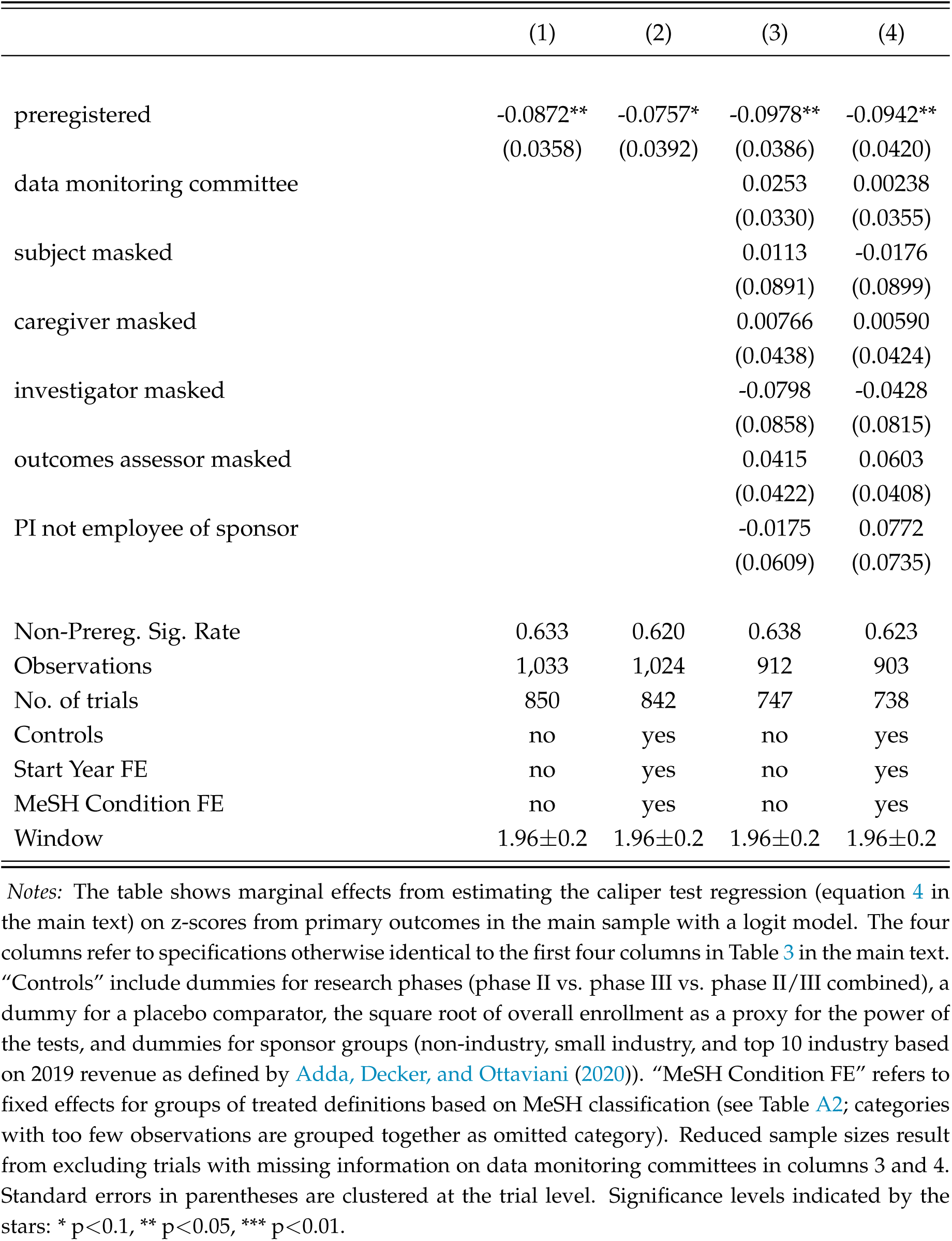
Caliper Tests for *z ∈*1.96*±*0.2 (Primary Outcomes, Marginal Effects from Logit Model without Sponsor FE)

## Footnotes

1 See, for instance, Munafò et al. (2017) and Christensen and Miguel (2018).

2 DiMasi, Grabowski, and Hansen (2016) estimate from surveys of pharmaceutical companies an average out-of-pocket cost of US$1,395 million (2013 dollars) per approved new compound.

3 The methods applied in this paper, based on analyzing the distribution of reported p-values, should be interpreted as joint tests for p-hacking and/or selective reporting but cannot reliably distinguish these two closely related notions.

4 Another commonly raised concern about preregistration, which is beyond the scope of the analysis in this paper, is that tying researchers’ hands too much may be detrimental to knowledge creation in some cases. Mandating strict adherence to pre-specified plans and discounting non-preregistered findings too much may discourage researchers from exploring potentially interesting observations and new theories of which they did not think ex-ante, or from conducting certain experiments altogether. See Coffman and Niederle (2015) and Banerjee et al. (2020) for discussions of these issues and potential solutions.

5 Similar methods have been used to study p-hacking and publication bias in academic publications in different disciplines, for instance, in economics (Brodeur et al., 2016; Vivalt, 2019; Brodeur, Cook, and Heyes, 2020), political sciences (Gerber and Malhotra, 2008), life sciences (Holman et al., 2015), and psychology (Simonsohn, Nelson, and Simmons, 2014). Elliott, Kudrin, and Wüthrich (2022*a*) and Elliott, Kudrin, and Wüthrich (2022*b*) provide an overview and a theoretical examination of different methods to detect p-hacking.

6 Other recent contributions in economic theory attempt to formalize the impact of preregistration, modeling it as a commitment device in persuasion games between researchers and evaluators (Felgenhauer, 2021; Kasy and Spiess, 2022; Williams, 2022).

7 Moreover, we exclude 31 trials that report an implausibly high number of over 30 primary outcome measures, and we follow Adda, Decker, and Ottaviani (2020) to exclude 25 trials of the sponsor *Colgate Palmolive*, which reported p-values precisely equal to 0.05 for 137 out of its 150 results. We attributed this to a reporting mistake; clearly, these were intended to be reported as significant results with a p-value lower than or equal to 0.05. Leaving *Colgate Palmolive*’s results in the sample would lead to a substantial spike at *z* = 1.96, which could be wrongly interpreted as evidence for p-hacking.

8 32.4% of p-values from primary outcomes, which meet the other criteria to be included in our sample, are not reported precisely but only relative to some threshold; for example, *p <* 0.001 or *p >* 0.1. 89.9% of these cases are highly significant results which are commonly reported as *p <* 0.001 (corresponding to *z >* 3.29) and *p <* 0.0001 (corresponding to *z >* 3.89). As barely any p-values are reported with equality below these thresholds, we lack information about the shape of the very left tail of the p-distribution (and the right tail of the z-distribution). For other parts of the distribution, including the region around the 5% significance threshold we focus on, such relative reporting is very infrequent. Therefore, we are confident in considering only exactly reported p-values for our tests to detect irregularities in the distribution at the 5% significance threshold.

9 Note that most p-values do not *actually* originate from a two-sided Z-test but from various other statistical procedures.

10 These criteria generally include that the study is interventional, concerns an FDA-regulated product, is not a phase I or feasibility study, and trial or manufacturing sites are located in the United States. See Zarin et al. (2016) and https://clinicaltrials.gov/ct2/manage-recs/fdaaa (accessed on April 5, 2023) for further details.

11 The *start date* of a trial refers to the date on which the first participant was enrolled in the clinical study.

12 The *primary completion date* of a trial refers to the date the last participant in a clinical study was examined or received an intervention to collect final data regarding one of the primary outcome measures.

13 While Table 1 only considers the trials that report at least one exact p-value (the main sample of our analysis, red ellipse in Figure 1), Appendix Table A3 provides a similar comparison for all applicable trials in the registry (blue ellipse in Figure 1). The correlation patterns are very similar.

14 For simplicity, we will refer to data monitoring committees, blinding, and independent PIs as “superior” design features throughout the paper because, if feasible to implement, these features are commonly considered superior regarding research integrity. However, we acknowledge that certain constraints may preclude implementing some of these features in some cases. Therefore, a trial not having all these features is not necessarily proof of inferior research integrity.

15 The FDAAA specifies fines for non-compliance with reporting requirements of over US$10,000per day. However, these regulations have not been enforced for a long time (Piller, 2020). The first notice of non-compliance was issued only in April 2021. As of April 2023, only four such notices were issued, none of which has led to an actual civil money penalty so far (see https://www.fda.gov/science-research/fdas-role-clinicaltrialsgov-information/clinicaltrialsgov-notices-noncompliance-and-civil-money-penalty-actions, accessed on April 8, 2023).

16 See, for instance, Anderson et al. (2015), Zarin et al. (2017), and DeVito, Bacon, and Goldacre (2020) for studies in the medical literature that evaluate compliance with registration and result reporting requirements to *ClinicalTrials.gov*. Adda, Decker, and Ottaviani (2020, Online Supplement) and Abrams, Libgober, and List (2023, Appendix G) provide more detailed reviews of this literature.

17 Intuitively, density discontinuity tests can be seen as the limit of caliper tests when the bandwidth *h* approaches zero.

18 Unfortunately, *ClinicalTrials.gov* only contains limited information about the actual researchers involved in conducting a study. Therefore, an analysis with researcher fixed effects is not feasible.

19 See, for instance, Ofosu and Posner (2020) and Banerjee et al. (2020) for discussions of these issues and potential solutions to them.

